# The Triangulation WIthin A STudy (TWIST) framework for causal inference within Pharmacogenetic research

**DOI:** 10.1101/2021.05.04.21256612

**Authors:** Jack Bowden, Luke C. Pilling, Deniz Türkmen, Chia-Ling Kuo, David Melzer

## Abstract

In this paper we review the methodological underpinnings of the general pharmacogenetic approach for uncovering genetically-driven treatment effect heterogeneity. This typically utilises only individuals who are treated and relies on fairly strong baseline assumptions to estimate what we term the ‘genetically moderated treatment effect’ (GMTE). When these assumptions are seriously violated, we show that a robust but less efficient estimate of the GMTE that incorporates information on the population of untreated individuals can instead be used. In cases of partial violation, we clarify when Mendelian randomization and a modified confounder adjustment method can also yield consistent estimates for the GMTE. A decision framework is then described to decide when a particular estimation strategy is most appropriate and how specific estimators can be combined to further improve efficiency. Triangulation of evidence from different data sources, each with their inherent biases and limitations, is becoming a well established principle for strengthening causal analysis. We call our framework ‘**T**riangulation **WI**thin a **ST**udy’ (TWIST)’ in order to emphasise that an analysis in this spirit is also possible within a single data set, using causal estimates that are approximately uncorrelated, but reliant on different sets of assumptions. We illustrate these approaches by re-analysing primary-care-linked UK Biobank data relating to *CYP2C19* genetic variants, Clopidogrel use and stroke risk, and data relating to *APOE* genetic variants, statin use and Coronary Artery Disease.

## 1 Background

Over the last 20 years the field of Epidemiology has embraced the exploitation of random genetic inheritance to help uncover causal mechanisms of disease using the technique of Mendelian randomization (MR). [1]. The basic premise of MR is illustrated by the causal diagrams in Fig 1: Genetic variants, usually Single Nucleotide Polymorphisms or SNPs, *G*, are found which robustly explain variation in a modifiable risk factor, *X*, where *X* is typically continuous (for example a person’s body mass index). The association between the exposure and an outcome, *Y*, hypothesised to be a downstream consequence of *X*, may be contributed to in observational data by unobserved confounding, *U*. If present, such confounding would mean that the naive association between *X* and *Y* would not reflect the causal effect of *X* on *Y*. If the important confounders could be appropriately measured and adjusted for, and no systematic selection bias or loss to follow up was present in the data (an assumption we will take as given for the remainder of the paper), then individuals with the same exposure level would be exchangeable [2] and observational associations could be interpreted causally. An MR analysis aims to circumvent any potential confounding by instead measuring the association between the outcome and the portion of the exposure that can be genetically predicted by the SNPs. Provided that the SNPs are independent of the confounders, are not associated with the outcome through any other pathway except the exposure, and the causal effect of a unit increase in the exposure is the same across individuals, then MR can consistently estimate the average causal effect of intervening on the exposure (e.g. to reduce or increase it) on the outcome. The Instrumental Variable (IV) assumptions of the MR approach are denoted IV1-IV4 in Fig 1.

**Fig 1:**
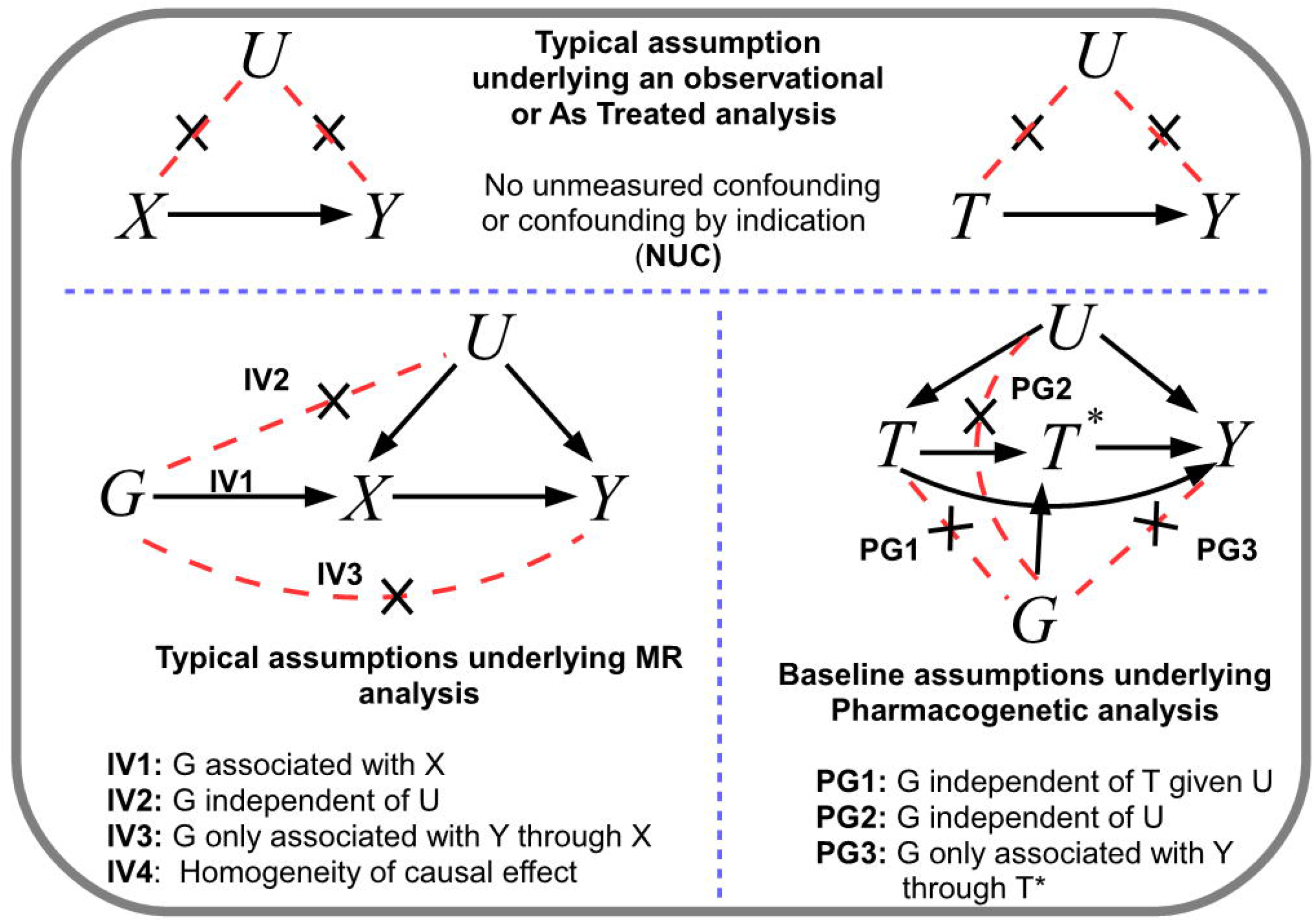
Top: Typical assumptions underlying the causal interpretation of an exposure *X* or treatment *T* on an outcome *Y* following standard regression analysis. Note that we assume NUC implies exchangeability : B = Bottom-left: Assumptions underlying a standard MR analysis using genetic variant G as an IV to estimate the causal effect of X on Y. C = Bottom-right: Assumptions underlying a standard pharmacogenetic analysis.

Genetic variants can also play an important role in helping to explain treatment effect heterogeneity in pharmacogenetics research. A canonical example is Clopidogrel: the primary drug for ischemic stroke prevention in the UK and many other countries (see https://cks.nice.org.uk/antiplatelet-treatment). It requires CYP2C19 enzyme activation in order to be properly metabolised into the active form of the drug and thus work to its fullest extent. However, it has long been known that both common loss-of-function and gain-of-function variants within the *CYP2C19* gene region can massively impact each patient’s ability to metabolise the drug [3]. Consequently, when prescribed in a primary care setting its effectiveness is heterogeneous, working for some and not for others. Estimating the true effectiveness of a treatment from observational data is challenging due to ‘confounding by indication’[4] (see Fig 1). For example, Clopidogrel use will, quite rightly, strongly depend on an individual’s underlying risk of stroke. However, unmeasured socio-economic factors may also influence both an individual’s ability to access appropriate healthcare and their underlying stroke risk [5]. Use of the drug for a sustained period may additionally depend on whether it can be tolerated without side-effects. Whilst these confounding factors could in principle be directly accounted for in a statistical analysis if complete information on a patient’s clinical state were available, this is seldom the case. A well known example (albeit in a non-pharmacogenetic context) where confounder adjustment failed is hormone replacement therapy, which was linked to increased cancer risk in observational data but not in subsequent randomized trials [6]. Thankfully, the need for adjustment can be circumvented if the purpose of the analysis is is instead to compare the relative effectiveness of a treatment across genetic groups (see Fig 1). In the case of Clopidogrel and *CYP2C19*, typically one might assume that *CYPC219* variants *G*: do not predict whether an individual receives Clopidogrel *T*; are not associated with any confounders predicting Clopidogrel use and stroke, *Y*; and only affects stroke risk through their interaction with Clopidogrel. We will refer to these as the ‘Pharmacogenetic’ (PG) assumptions as a counterpart to the IV assumptions utilised by MR. A key difference exists between the role of the gene in MR and the role of the gene in pharmacogenetics: In MR, genes are assumed to directly influence a modifiable exposure. In Pharmacogenetics we can think of treatment as the exposure, and the genes are hypothesised to alter treatment *once* it is taken. In Fig 1 we denote the genetically altered treatment with the symbol *T \**.

In recent work, Pilling et al [7] use data from GP-linked UK Biobank participants on Clopidogrel treatment to estimate its effect in different *CYP2C19* genetic subgroups and from this the number of strokes that could potentially be avoided if all individuals could experience the same benefit as the group with the most favourable genotype (through either dose modification or through switching to an alternative drug). In this paper we review the methodological underpinnings of the general PG approach, which utilises only individuals who are treated and relies on fairly strong baseline PG assumptions to estimate what we refer to as the ‘Genetically Moderated Treatment Effect’ (GMTE). When the PG assumptions are violated, we show that a robust but less efficient estimate of the GMTE that incorporates information on the population of untreated individuals can instead be used. In cases of partial violation, we clarify when Mendelian randomization and traditional confounder adjustment can also yield consistent estimates for the GMTE. A decision framework is then described to decide when a particular estimation strategy is most appropriate and how specific estimators can be combined to further improve efficiency.

Triangulation of evidence from different data sources, each with their inherent biases and limitations, is becoming a well established principle for strengthening causal analysis [8]. We call this framework ‘**T**riangulation **WI**thin a **ST**udy’ (TWIST)’ in order to emphasise that an analysis in this spirit is also possible within a single data set, using causal estimates that are approximately uncorrelated and reliant on different sets of assumptions. This makes their estimates easy to quantitatively combine if sufficiently similar to improve the precision and robustness of any findings. More broadly, it enables estimates to be qualitatively compared and contrasted, with expert judgement used to assess whether their assumptions are likely to have been met, in order to come to an overall conclusion about the totality of evidence. We illustrate these approaches by re-analysing primary-care-linked UK Biobank data relating to *CYP2C19* genetic variants, Clopidogrel use and stroke risk, and data relating to *APOE* genetic variants, statin use and Coronary Artery Disease (CAD).

## 2 Methods

Suppose that we are interested in evaluating the maximal effectiveness of a treatment *T* on an outcome *Y* using observational data. For simplicity we will assume initially that *T* is a binary treatment indicator so that, if prescribed, it is taken in full, that *Y* is a continuous or binary outcome variable and we are interested in estimating the treatment effect as a mean or risk difference contrast. A simple but naive way of estimating this effect would be to compare outcomes across those who are treated and those who are untreated. Borrowing terminology from the clinical trials literature, we refer to this as the ‘As treated’ (AT) estimate (Fig 1 top right). The AT estimate may not directly address our research needs for two reasons. The first reason is that, although we may understand many of the factors which influence whether an eligible individual is prescribed treatment by their doctor, there may be unmeasured variables, *U*, which influence both the decision to prescribe treatment and the outcome. Indeed, even if the treatment is truly effective in reducing the severity or risk of *Y*, it is highly likely that the population of treated individuals may still experience worse outcomes than those who are untreated. This would mean that the sign of the AT estimate could be positive, and thus qualitatively different than the true causal effect. This is classic confounding by indication. The second reason is that a pharmacogenetic investigation may suggest that the treatment does not in fact work for a certain proportion of the population at all, has a markedly reduced effectiveness, or increases the risk of side-effects.

We would like to estimate the difference in patient outcomes if all patients who take treatment experienced the ‘full’ effect, as experienced by those with the treatment-enabling genotype versus the reduced (or zero) effect experienced by those with a treatment-inhibiting genotype. To realise such a benefit in practice, we could switch patients with the treatment-inhibiting genotype to an alternative medication which then works to its fullest extent. We will call this hypothetical quantity (or ‘estimand’) the Genetically Moderated Treatment Effect (GMTE).

### 2.1 The causal estimand and key identifying assumptions

To make the target of our analysis more explicit we will assume a simple model with a binary genotype *G*, where *G*=1 denotes the treatment-enabling genotype and *G*=0 denotes the treatment-inhibiting genotype. We now define a new treatment-moderating variable *T* ^*\**^ which is equal to the product or interaction *T* × *G*. We consider the following simple linear interaction model for the expectation (or mean) of outcome *Y* given treatment, *T*, genetic variant *G*, measured confounder *Z* and unmeasured confounder *U* :

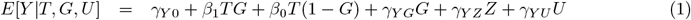

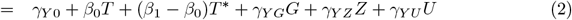

Under model (1), *β*_1_ and *β*_0_ reflect the treatment effect experienced by those with genotype *G*=1 and *G*=0 respectively and thus allows for genetically driven treatment effect heterogeneity. The parameter *γ*_*Y G*_ represents the direct effect of *G* on *Y* and *γ*_*Y U*_ represents the direct effect of *U* on *Y*. To clarify the causal estimand of interest we re-write model (1) as model (2). Using potential outcomes notation, we can express the GMTE estimand as the average causal effect if everyone could receive moderated treatment level *T*\*=1 (i.e. the full or enhanced effect) versus if everyone could receive treatment level *T*\*=0 (i.e. no enhanced effect):

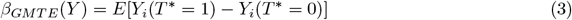

This is equal to *β*_1_ − *β*_0_, the coefficient of *T*\* in model (2). We now define the key assumptions that will be leveraged by the various methods proposed in this paper. These assumptions are also represented by the causal diagram and corresponding association parameters in Fig 2:

**Fig 2:**
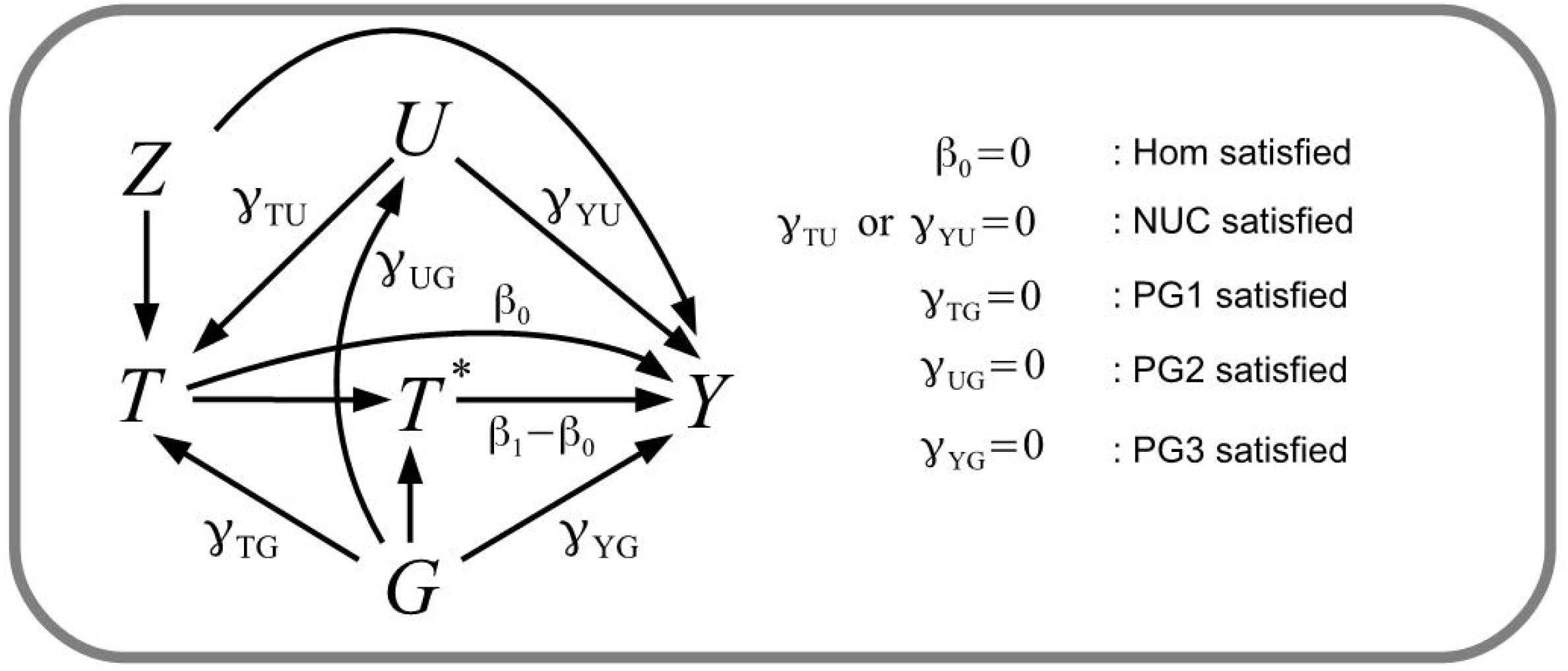
Causal diagram explaining the key assumptions leveraged by the methods proposed. The diagram and notation are consistent with outcome model (2) and the simulation simulation study

- **Homogeneity (Hom)**: Individuals who take treatment with genotype level *G*=0 experience no treatment effect all (*β*_0_ = 0). Note that this is subtly different to Homogeneity assumption IV4 made in Mendelian randomization, which in this context would state that *β*_1_ = *β*_0_;
- **PG1**: An individual’s genotype *G* is independent of the decision to take treatment, *T*, given all confounders, *U*, of *T* and outcome *Y* (*γ*_*T G*_=0);
- **PG2**: An individual’s genotype *G* is independent of confounders *U* (*γ*_*UG*_=0);
- **PG3**: An individual’s genotype *G* is independent of their outcome *Y* given treatment *T* and all confounders *U* (*γ*_*Y G*_=0);
- **No unmeasured confounding (NUC)**: All confounding variables *U* that predict *T* and *Y* have been measured and adjusted for (*γ*_*Y U*_ or *γ*_*T U*_ =0).

As previously stated, we will assume that the NUC assumption implies exchangeability between treatment groups, which rules out the presence of systematic selection bias or systematic loss to follow up in the data.

### 2.2 Estimating the GMTE by correcting the As-Treated estimate

The As-Treated estimate suffers in general from confounding by indication, and a lack of specificity to the genetic variant driving the mechanistic interaction with the treatment. However, if the Hom and NUC assumptions are satisfied and either PG1 or PG3 are satisfied, then, the ‘Corrected’ As Treated estimate (CAT) can consistently estimate the GMTE. Put simply, if confounding bias can be addressed and the population treatment effect is driven entirely by the *G*=1 subgroup, then the correct quantity can be estimated by scaling the treatment-outcome association by the proportion of treated individuals with the *G*=1 genotype.

### 2.3 Estimating the GMTE in the treated population only

We next consider estimation of the GMTE in the general case where only assumptions PG1-PG3 hold. Together they imply that *G* is jointly independent of treatment and any unmeasured confounders, and they only affect the outcome through the treatment moderator variable *T*\*. Among the population of treated individuals, we can think of an individual’s genotype as randomly allocating them to either moderated treatment level *T*\*=1 or *T*\*=0. This means we can calculate the GMTE using only treated individuals via the ‘GMTE(1)’ estimate. We use the additional subscript ‘(1)’ in the estimate’s nomenclature to denote that it conditions on *T* =1.

### 2.4 A robust GMTE estimator

We next consider estimation of the GMTE under violations of PG1-PG3. Violation of PG1 implies that an individual’s genotype directly influences the likelihood that they receive treatment. For example, it could be that those with a *G*=0 genotype have an increased risk of side effects on treatment and choose to immediately come off the drug. An alternative explanation could be genetic population stratification [9]: e.g the allele frequency of the genetic variant *G* and the rate of treatment could simultaneously vary across individuals from different ethnic groups. An example of PG2 violation would be if the genetic variant increases the likelihood of an unmeasured risk factor for the outcome, and this risk factor also increases their likelihood of being treated. An example of PG3 violation would be if an individual’s genotype directly affects the outcome through a pathway completely independent of either treatment or any confounding factor, which could be viewed as horizontal pleiotropy [10]. When any of these assumptions are violated the GMTE(1) estimate will reflect the genetically moderated effect of treatment plus the bias due to PG1-3 violation. Specifically, this bias would be the sum of:

- *b*_*PG*1_ via the *G* → *T* ← *U* → *Y* pathway (due to violation of PG1);
- *b*_*PG*2_ via the *G* → *U* → *Y* pathway (due to violation of PG2);
- *b*_*PG*3_ via the *G* → *Y* pathway (due to violation of PG3).

Whilst the bias contributions *b*_*P G*2_ and *b*_*P G*3_ are clear, bias contribution *b*_*P G*1_ is perhaps less so; it occurs because the GMTE(1) estimate explicitly conditions on treatment *T*, the presence of an association between *G* and *T* makes *T* a ‘collider’ [11]. This is one example of broader point: the RGMTE estimate is not robust to effect modification of any variable associated with T. Thankfully, when assumption PG1 is satisfied and no other effect modification is present, bias terms *b*_*P G*2_ and *b*_*P G*3_ can be consistently estimated and removed by incorporating information on the untreated population. This is achieved by calculating the equivalent GMTE(1) estimate for the untreated group and then subtracting this estimate from that in the treated group. We call this the ‘Robust’ genetically moderated treatment effect (RGMTE) estimate. Although assumption PG1 is key for the RGMTE estimate, we show that it can work if PG1 is violated but the NUC assumption is satisfied.

### 2.5 A ‘Mendelian randomization’ estimate

Given data on both treated and untreated individuals, it is possible to obtain an estimate for the GMTE by using the genetic variant *G* as an instrumental variable for the treatment moderator variable *T*\* directly, as in Mendelian randomization. In the context of a single gene, *G*, the MR estimate is the ratio of the gene-outcome association and the the gene-*T*\* association. The MR estimate is consistent for the GMTE if PG2-PG3 hold and either PG1 holds, or the Hom assumption holds.

In *S1 Text* we provide a formal justification of when the CAT, GMTE(1), RGMTE and MR estimates are consistent for the GMTE assuming outcome model (2)..

### 2.6 Method summary and implementation

In Table 1 we: (i) give statistical formulae for the GMTE(1), GMTE(0), RGMTE, MR and CAT estimates; (ii) provide more detailed information on the sufficient assumptions each one relies upon to consistently estimate the GMTE (or in the case of the GMTE(0) estimate, zero); (iii) show how to test whether potential confounders of treatment and outcome could bias each estimate; and (iv) give generic R psuedocode to obtain each estimate. To further clarify point (iii), take the GMTE(1) estimate as an example. In order to assess whether a potential confounder could meaningfully bias its estimate, we calculate the GMTE(1) estimate but use a potential confounder *U* as the outcome in place of the true outcome *Y*. If this GMTE(1) estimate is significantly non-zero then it indicates a meaningful bias in the GMTE estimate with respect to the outcome, unless the confounder is adjusted for by treating it as an additional component of *Z*. This principle holds for all other GMTE estimates as well. Unlike the GMTE(1) and RGMTE estimators, the MR and CAT estimators both have a ratio form with the denominator dependent on *G*. For this reason they are more susceptible to bias and imprecision when the sample size is small and *G* has a low allele frequency.

**Table 1:**
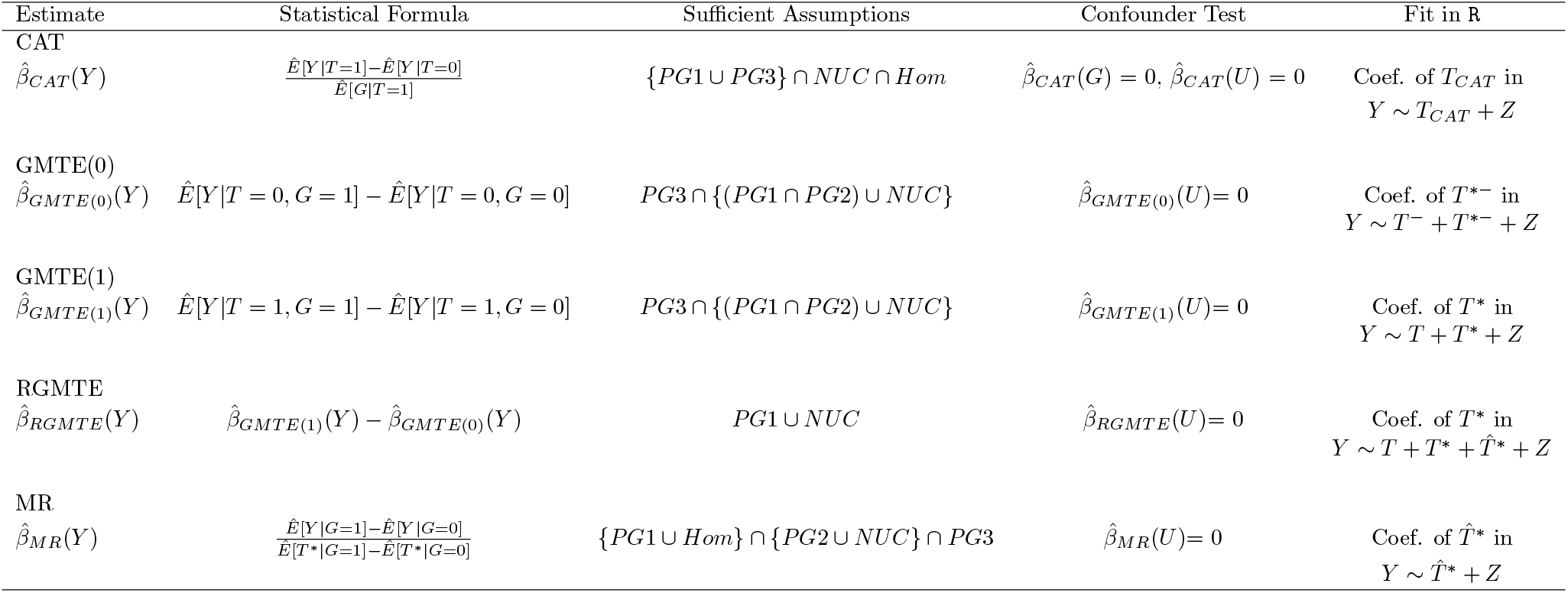
Columns left to right show: Statistical formulae for GMTE(1), GMTE(0), RGMTE, MR and CAT estimates; Sufficient assumptions each one relies upon to consistently estimate the GMTE (or zero in the case of the GMTE(0) estimate); Estimate-specific confounder test statistics; generic R code to obtain each estimate. For the GMTE(0) estimate, 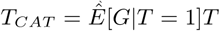, for the GMTE(0) estimate *T* ^*−*^ = 1 *− T, T* ^*\*−*^ = *T* ^*−*^*G*, for the RGMTE estimate *T*\* = *T G* and 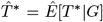. Note that the GMTE(0) estimate does not directly target the GMTE, but rather zero under the PG assumptions..

When the outcome is continuous, the approaches can be implemented using linear regression to estimate the GMTE as a mean difference. With a binary outcome, we recommend estimating risk differences directly using either a linear probability model, or using a logistic regression model to furnish estimates on the risk difference scale. With time-to-event data, we recommend analysing the data under an additive hazards model. Further details are provided *S1 Text*. We suggest to estimate mean difference, risk differences or additive hazard differences in order to guarantee that we can obtain estimates for the GMTE from different estimators on the same scale, because these measures are collapsible. That is, they should remain constant when marginalised over unobserved confounders [12]. This is especially important for being able to effectively combine methods, as described in the next section.

### 2.7 Which estimates can be combined?

When two estimates are highly correlated, we gain little knowledge when they are observed to be similar. However, when two uncorrelated or weakly correlated estimates are similar, it gives credence to the hypothesis that they are estimating the same underlying quantity, and there is the potential to combine them into a single, more precise estimate. In *S1 Text* we show that the RGMTE and MR estimates are asymptotically uncorrelated. We also show that the CAT estimate is mutually uncorrelated with the GMTE(1) and RGMTE estimates, and uncorrelated with the MR estimate when *G* is independent of *T*. In cases where *G* and *T* are not perfectly independent, but *G* is a modest predictor of *T* (a highly plausible scenario in most pharmcogenetic contexts), the correlation between the MR and CAT estimate will be non-zero but practically negligible. The fact that most estimate-pairs are uncorrelated makes them easy to combine via a simple inverse variance weighted average or meta-analysis. In order to decide whether two uncorrelated estimates can be combined, we propose the use of a simple heterogeneity statistic. This procedure is illustrated in Fig 3 taking the GMTE(1) and CAT estimates as an example. Using each estimate we calculate their inverse variance weighted average and from this the heterogeneity statistic, *Q*_*GMT E*(1),*CAT*_. If this statistic is less than the 1-*α* quantile of a 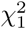 density (where *α* is the pre-specified significance threshold) then we judge the GMTE(1) and CAT estimates to be sufficiently similar to combine into a single estimate more efficient estimate. If *Q*_*GMT E*(1),*CAT*_ is greater than 1-*α* threshold then the two estimates should be left separate. Along with single estimates, combined estimates that meet this heterogeneity statistic are colour coded blue (e.g. Fig 3 case (i)) and those which do not will be colour coded black (e.g. Fig 3 case (ii)). We stick to this convention for the remainder of the paper.

**Fig 3:**
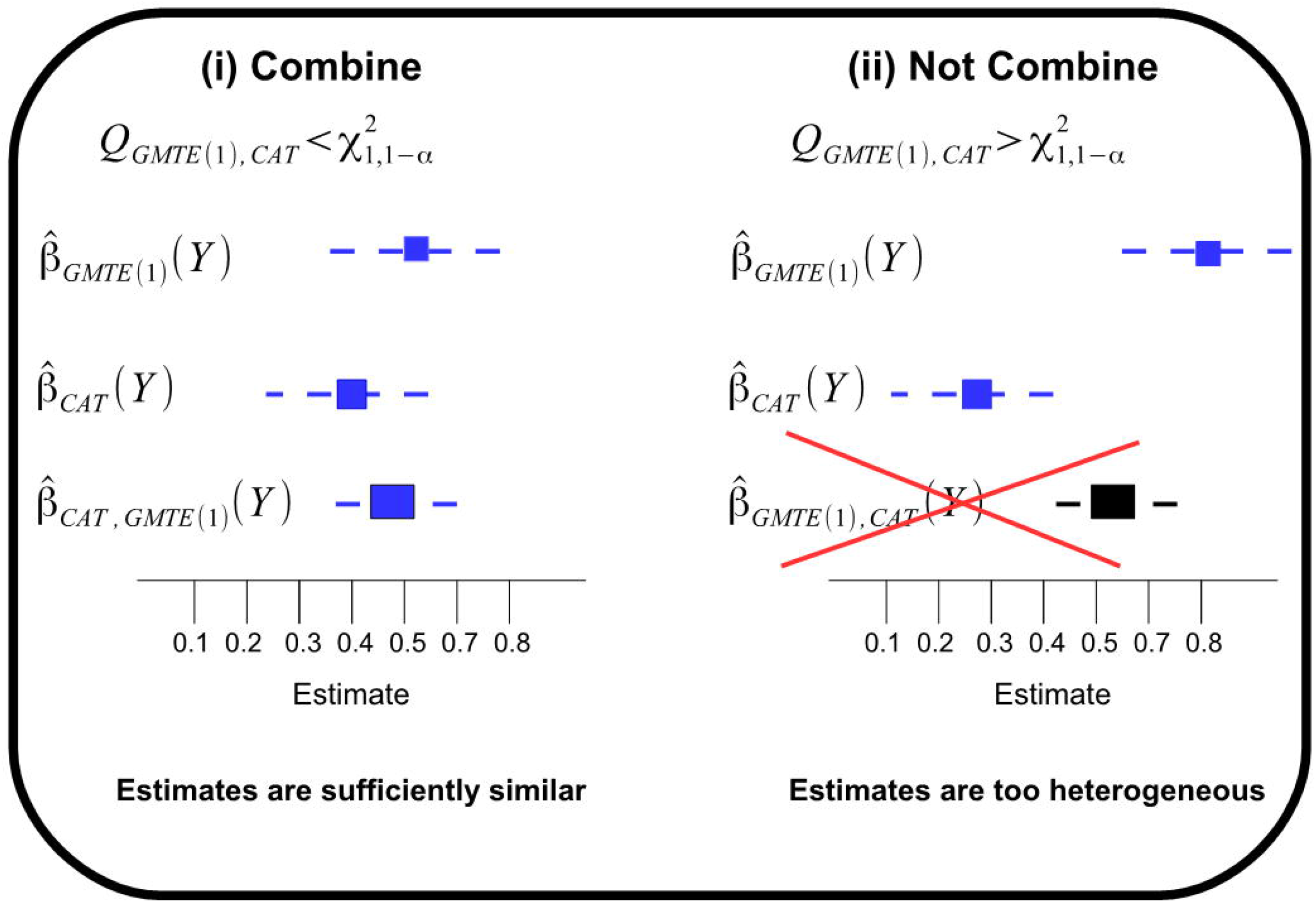
Two statistically uncorrelated estimates are homogenous enough to be meaningfully combined - case (i) - or are too heterogeneous to be combined - case (ii)

The RMGTE and MR estimates are in general highly correlated with the GMTE(1) estimate, and should therefore not be combined. *S1 Text* we show that the MR and RMGTE estimates can be viewed as complementary functions of the GMTE(1) and GMTE(0) estimates, and that the combined RGMTE/MR estimate is exactly equivalent to the GMTE(1) estimate when *G* is independent of *T* and the proportion of treated and untreated participants in the data is the same. Fig 4 shows a pictorial diagram of all single and combined estimates that can be derived using the above heterogeneity statistic criteria. This comprises four original estimates (CAT,GMTE1,RMGTE,MR), four ‘paired estimates (CAT/GMTE1, CAT/RGMTE, CAT/MR, RGMTE/MR) and one ‘triplet’ estimate (CAT/RGMTE/MR), making nine in total. One possible analysis option would be to report all single and valid combined estimates which are sufficiently homogeneous according to a particular significance threshold. Another option would be to allow the GMTE(0) estimate to initially guide the analysis towards either the GMTE(1) estimate (and its possible combination with the CAT estimate) or the RMGTE estimate (and its possible combination with either MR estimate, the CAT estimates or both). Alternatively, some may be more comfortable with a qualitative assessment of the totality of evidence gleaned across the four distinct analysis procedures, using prior scientific knowledge to weigh up their individual importance after careful consideration as to the plausibility of their key assumptions.

**Fig 4:**
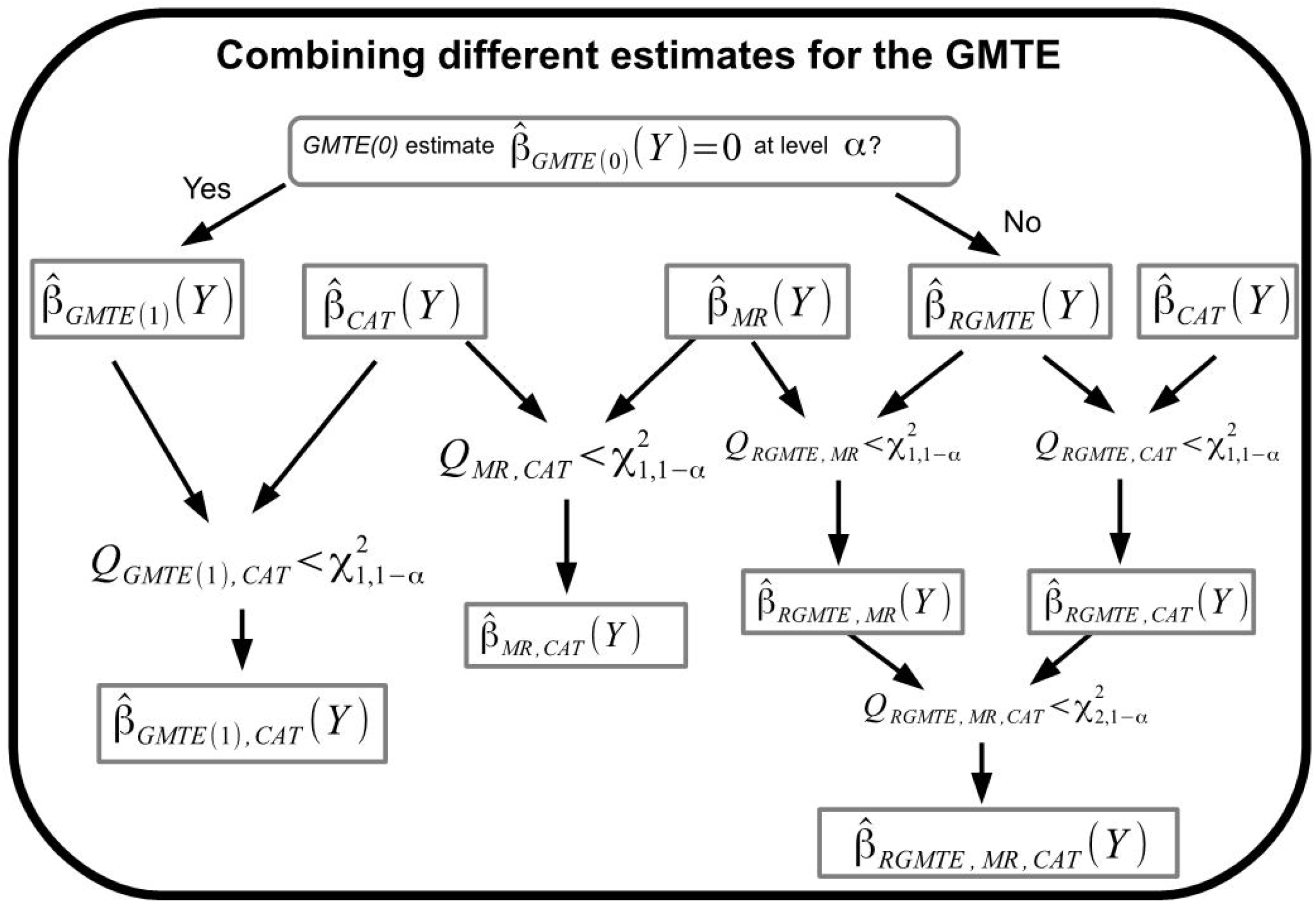
A schematic diagram showing all possible 9 single, two-way or three-way combined estimators of the GMTE that can be calculated using the TWIST framework.

## 3 Simulation illustration

Trial data comprising a binary genotype *G*, treatment indicator *T*, observed covariate *Z* and a continuous outcome *Y* are simulated for *n*=10,000 patients using the following data generating model which is consistent with the causal diagram in Fig 2:

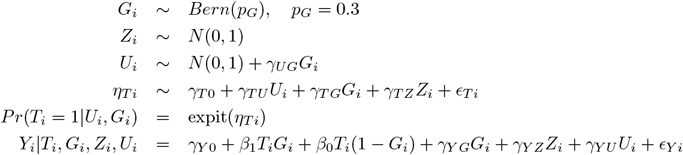

Under this model, assumptions PG1-PG3 are violated if *γ*_*T G*_, *γ*_*UG*_ and *γ*_*Y G*_ are non-zero respectively. The Hom assumption is violated when *β*_0_. Finally the NUC assumption is violated if either *γ*_*T U*_ or *γ*_*Y U*_ (or both) are non-zero. For simplicity we keep *γ*_*Y U*_ fixed and non-zero and vary only *γ*_*T U*_. Note that if the NUC assumption holds, then PG2 is in a sense automatically satisfied because *U* is no longer a confounder. However, in this case there may still be a path from *G* to *Y* via *U*. This would then form all or part of any PG3 violation.

Fig 5 shows the distribution of estimates for the GMTE obtained across 500 independent data sets and six simulation scenarios, using the CAT, GMTE(1), RGMTE and MR estimators. We also show the distribution of the GMTE(0) estimate in each case, as a helpful guide to understand the extent of bias that can be estimated from the data. In all scenarios the true GMTE is fixed at -0.5. Table 2 shows the mean point estimates, standard errors and 95% confidence interval coverage corresponding to the same six scenarios. For the five combined estimators, Table 3 shows: mean point estimates, mean standard errors, 95% confidence interval coverage and the proportion of times each combined estimator passes the heterogeneity test using a significance threshold of *α*=0.05.

**Table 2:**
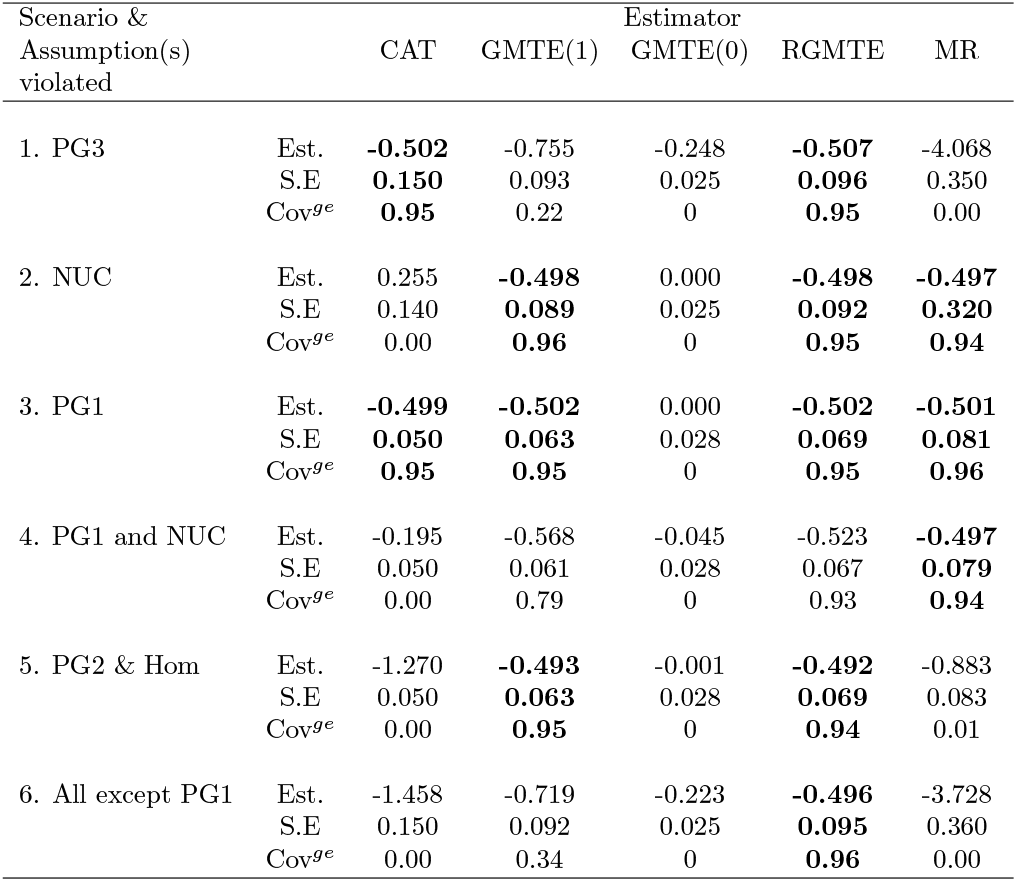
Mean point estimates, standard errors and coverage (of 95% confidence interval) for the CAT, GMTE(1), GMTE(0) RGMTE and MR estimates across six simulation scenarios. In each case, the true GMTE is fixed at -0.5. Unbiased estimates and associated standard errors/coverages are highlighted in bold.

**Table 3:**
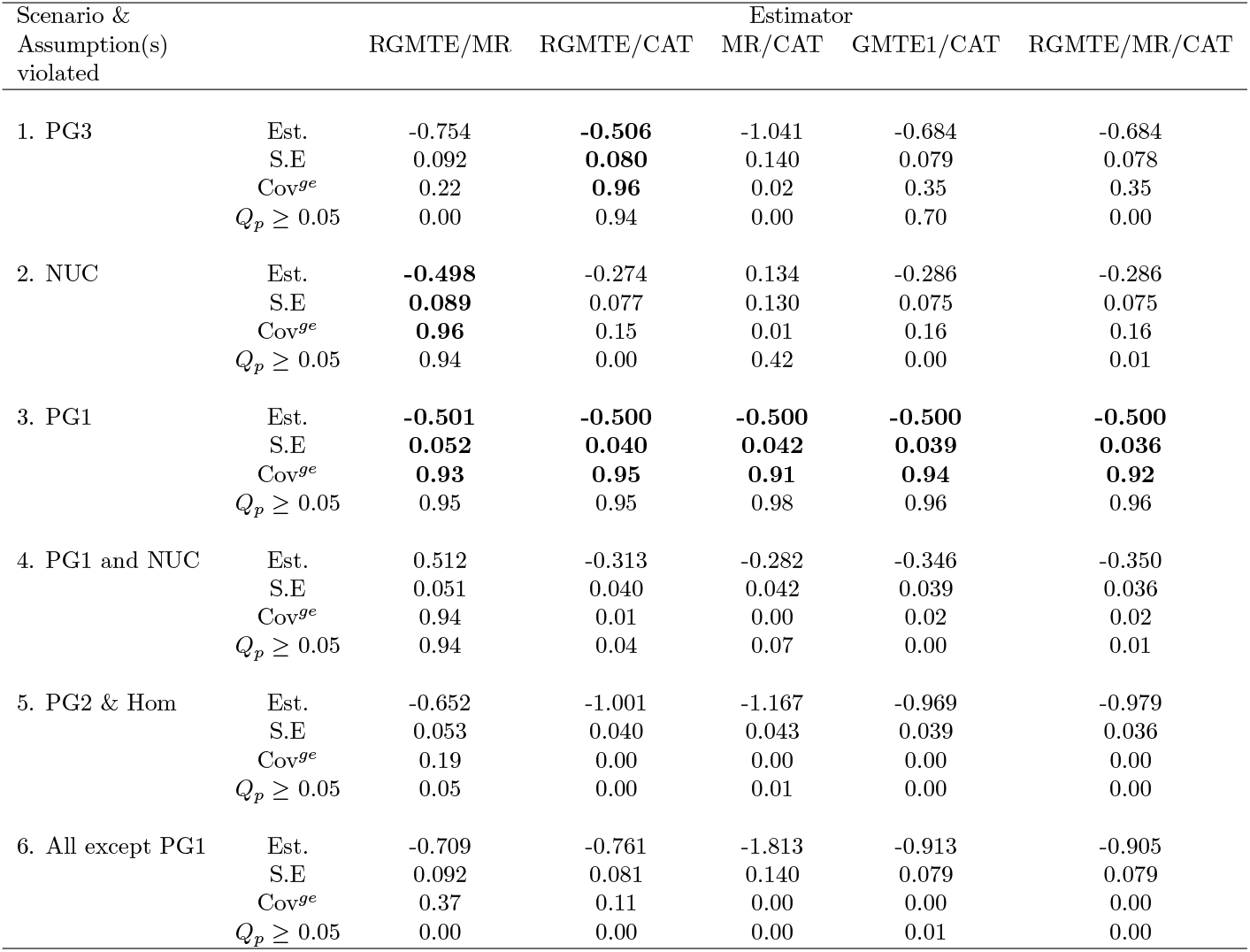
Mean point estimates, standard errors, coverage (of 95% confidence interval) and heterogeneity test rejection rates for the five combined estimates across six simulation scenarios. In each case, the true GMTE is fixed at -0.5. Unbiased estimates and associated standard errors/coverage are highlighted in bold.

| Scenario &<br>Assumption(s)<br>violated |  | Estimator |  |  |  |  |
| --- | --- | --- | --- | --- | --- | --- |
|  |  | RGMTE/MR | RGMTE/CAT | MR/CAT | GMTE1/CAT | RGMTE/MR/CAT |
| 1. PG3 | Est. | -0.754 | **-0.506** | -1.041 | -0.684 | -0.684 |
|  | S.E | 0.092 | **0.080** | 0.140 | 0.079 | 0.078 |
|  | Cov<sup>ge</sup> | 0.22 | **0.96** | 0.02 | 0.35 | 0.35 |
| | $Q_p \geq 0.05$ | 0.00 | 0.94 | 0.00 | 0.70 | 0.00 |
| 2. NUC | Est. | **-0.498** | -0.274 | 0.134 | -0.286 | -0.286 |
|  | S.E | **0.089** | 0.077 | 0.130 | 0.075 | 0.075 |
|  | Cov<sup>ge</sup> | **0.96** | 0.15 | 0.01 | 0.16 | 0.16 |
| | $Q_p \geq 0.05$ | 0.94 | 0.00 | 0.42 | 0.00 | 0.01 |
| 3. PG1 | Est. | **-0.501** | **-0.500** | **-0.500** | **-0.500** | **-0.500** |
|  | S.E | **0.052** | **0.040** | **0.042** | **0.039** | **0.036** |
|  | Cov<sup>ge</sup> | **0.93** | **0.95** | **0.91** | **0.94** | **0.92** |
| | $Q_p \geq 0.05$ | 0.95 | 0.95 | 0.98 | 0.96 | 0.96 |
| 4. PG1 and NUC | Est. | 0.512 | -0.313 | -0.282 | -0.346 | -0.350 |
|  | S.E | 0.051 | 0.040 | 0.042 | 0.039 | 0.036 |
|  | Cov<sup>ge</sup> | 0.94 | 0.01 | 0.00 | 0.02 | 0.02 |
| | $Q_p \geq 0.05$ | 0.94 | 0.04 | 0.07 | 0.00 | 0.01 |
| 5. PG2 & Hom | Est. | -0.652 | -1.001 | -1.167 | -0.969 | -0.979 |
|  | S.E | 0.053 | 0.040 | 0.043 | 0.039 | 0.036 |
|  | Cov<sup>ge</sup> | 0.19 | 0.00 | 0.00 | 0.00 | 0.00 |
| | $Q_p \geq 0.05$ | 0.05 | 0.00 | 0.01 | 0.00 | 0.00 |
| 6. All except PG1 | Est. | -0.709 | -0.761 | -1.813 | -0.913 | -0.905 |
|  | S.E | 0.092 | 0.081 | 0.140 | 0.079 | 0.079 |
|  | Cov<sup>ge</sup> | 0.37 | 0.11 | 0.00 | 0.00 | 0.00 |
| | $Q_p \geq 0.05$ | 0.00 | 0.00 | 0.00 | 0.01 | 0.00 |

**Fig 5:**
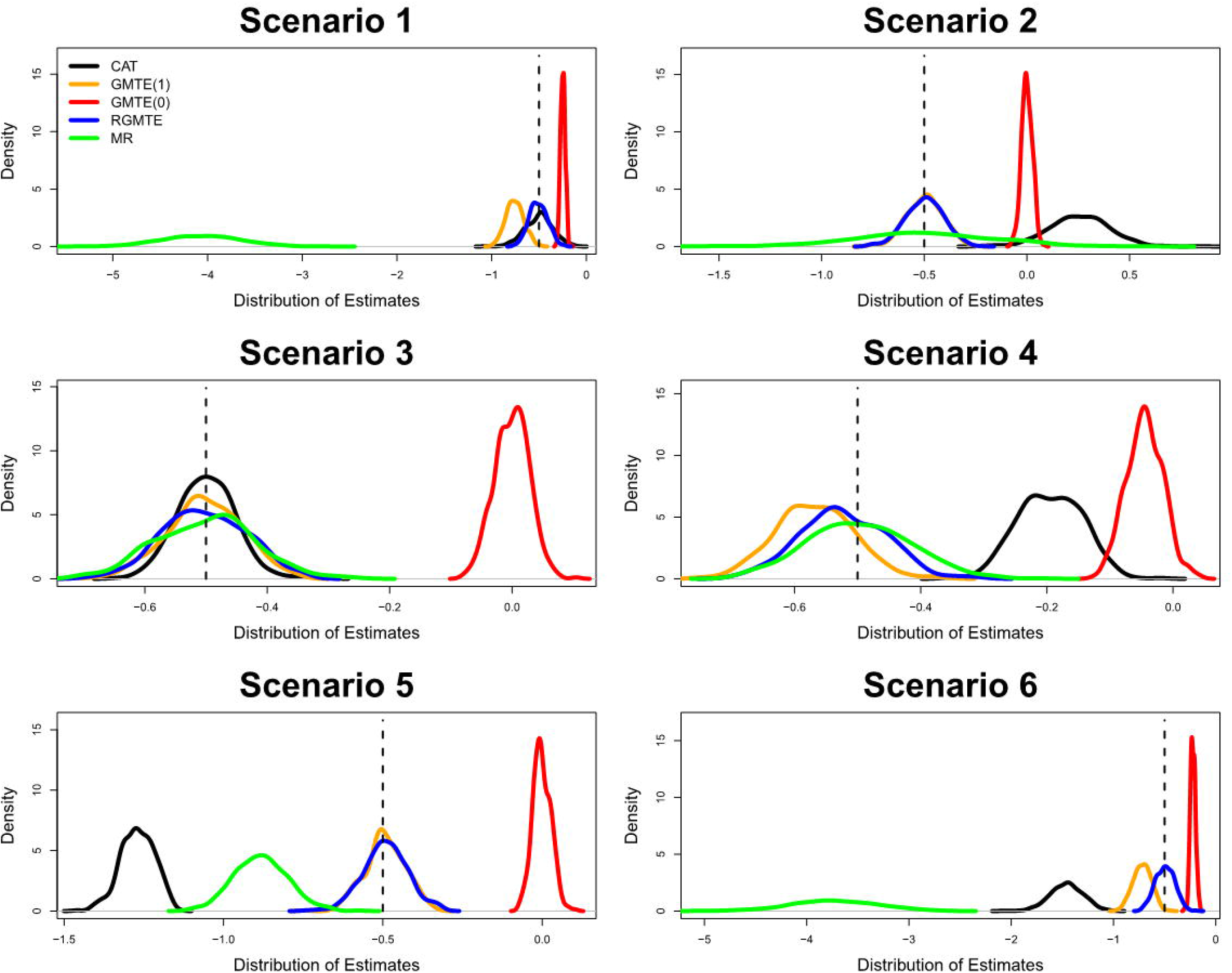
Distribution of estimates for the CAT, GMTE(1), GMTE(0), RGMTE and MR estimators across six simulation scenarios. In each case, the true GMTE is fixed at -0.5.

In Scenario 1 assumption PG3 is violated but all others (PG1, PG2, Hom, NUC) are satisfied. In this case both the CAT and RMGTE estimators are unbiased, with the RGMTE having the smallest standard error. In Table 3 we see that the combined RGMTE/CAT estimate is consequently unbiased with a standard error of 0.085, which is smaller than either the RMGTE or CAT estimates.

In Scenario 2 the NUC assumption is violated but all others (PG1, PG2, Hom) are satisfied. In this case the GMTE(1), RGMTE and MR estimators are unbiased, with the GMTE(1) estimate being the most precise. In Table 3 we see that the combined RGMTE/MR estimate is consequently unbiased with a standard error near-identical to the GMTE(1) estimate, in line with the theoretical prediction outlined in *S1 Text*. In Scenario 3 assumption PG1 is violated and (PG2, PG3, Hom, NUC) are satisfied. In this case all estimators are unbiased. In Table 3 we show in this case that the most efficient unbiased estimate of all comes from combining the RGMTE, CAT and MR estimates. In Scenario 4 PG1 and NUC are violated but the remaining assumptions (PG2, PG3, Hom) are satisfied. In this case only the MR estimate is unbiased. Consequently, no combined estimator is unbiased although the bias in the RGMTE/MR estimate is small. In Scenario 5, PG2 and Hom are satisfied but (PG1, PG3, NUC) are satisfied. In this case the GMTE(1) and RGMTE estimators are unbiased, with the GMTE(1) estimator being the most efficient. No combined estimate is unbiased. In Scenario 6 all assumptions except PG1 are violated. In this case only the RGMTE estimate is unbiased and, again, no combined estimate is unbiased.

In order to gauge the sensitivity of each estimator to the minor allele frequency of *G*, we repeat simulation scenario 3 for six values of *p*_*G*_ between 0.02 and 0.3. Fig 6 plots the mean standard error of the estimates in each case. We see clearly that the precision of all estimates is an increasing function of minor allele frequency. However, the loss in precision at low allele frequencies is strongest for the MR and CAT estimates. This is because they are both ratio estimates, with the denominator depending heavily on *G*.

**Fig 6:**
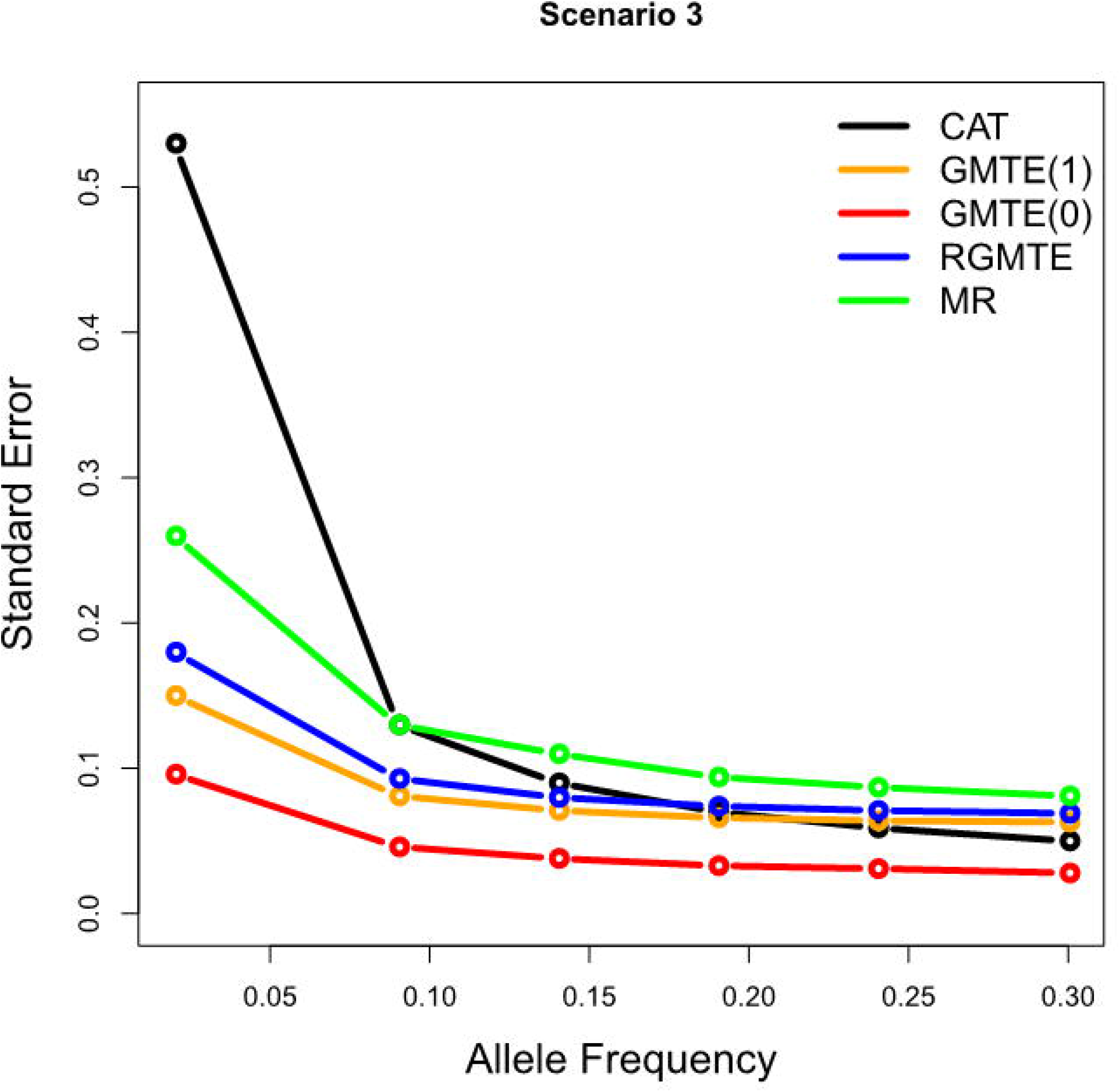
Mean standard error of the CAT, GMTE(1), GMTE(0), RGMTE and MR estimates for Scenario 3 as a function of the minor allele frequency of *G*

In conclusion, our simulation study provides an empirical verification of the strengths and limitations of each approach, and when any two uncorrelated estimates can be effectively combined via a simple inverse variance weighted meta-analysis. Although the standard error of any estimate that combines the CAT and MR estimates requires *G* to be independent of *T* to be strictly valid (since this implies a zero correlation between their estimates) when this assumption is violated in Scenario 3 it only induces a modest loss of coverage (e.g. 91% for the CAT/MR estimate and 92% for the CAT/RGMTE/MR estimate). Across the simulations the RGMTE is emerges as the most robust estimate.

## 4 Applied analyses

### 4.1 Clopidogrel, CYPC219 & Stroke risk

Clopidogrel is a widely used anti-platelet therapy that impairs platelet aggregation with consequent reductions in risk of atherothrombotic events such as myocardial infarctions and ischemic strokes [13]. Clopidogrel is a pro-drug that requires activation by liver enzymes, primarily CYP2C19. Genetic variants in *CYP2C19* impair function with subsequently reduced Clopidogrel active plasma levels [14], and we have previously shown using primary care linked data on UK Biobank participants that carriers of these variants have increased risks of ischemic stroke and myocardial infarction (MI) whilst prescribed Clopidogrel in the primary care setting [7]. In this work we calculated the population attributable fraction using established methods by analysing data on only those who were treated with Clopidogrel, but we revisit the analysis and apply the full decision framework proposed in this paper.

The UK Biobank study recruited 503,325 volunteers from the community who attended one of 22 assessment centres in England, Wales or Scotland between 2006 and 2010 [15]. Participants were aged 40 to 70 years at the time of assessment, and baseline assessment included extensive questionnaires on demographic, health, and lifestyle information. Blood samples were taken, allowing analysis of participant genetics. Ethical approval for the UK Biobank study was obtained from the North West Multi-Centre Research Ethics Committee. This research was conducted under UK Biobank application 14631 (PI: DM).

Linked electronic medical records from primary care are available for 230,096 (45.7%) of participants, which includes *>*57 million prescribing events between 1998 and 2017. Detailed description of the data extracted and limitations are available from UK Biobank. For this analysis we excluded 5,353 participants missing any genetics data, then 14,856 of non-European genetic ancestry, then 555 missing any *CYP2C19* loss of function genotype data, leaving 209,333 participants with sufficient primary care and genetic data. N=198,868 never received a Clopidogrel prescription. N=938 only ever received one prescription, so did not have sufficient exposure time for study. Of the 9,527 participants remaining, in 2,044 the prescribing frequency was less than once every 2 months, and these were also excluded. This left 7,483 participants with at least two Clopidogrel prescriptions for analysis. Baseline information on the included participants is shown in Table 4.

**Table 4:** Baseline data on UK Biobank participants in the Clopidogrel analysis set. *Based on hospital episode statistics data.

| Variable | Prescribed Clopidogrel<br>(n=7,483) | Never Prescribed Clopidogrel<br>n = 198,868 |
| --- | --- | --- |
| Age at recruitment | 61.4± 6.2 | 56.5±8.0 |
| Age at first prescription | 64.1±7.3 | - |
| Sex(Female%) | 2,559(34.2%) | 110,569(55.6%) |
| *CYP219* LoF carrier | 2,145(28.7%) | 56,043(28.2%) |
| Incident Ischemic Stroke diagnosis* | 110 (1.5%) | 2,078(1.0%) |
| Incident MI diagnosis* | 1,822 (24.8%) | 13,796(6.9%) |

*CYP2C19* loss-of-function (LoF) carriers (any*2-*8 alleles) had significantly increased ischemic stroke risk (Hazard Ratio (HR) 1.53: 95% CIs 1.04 to 2.26, p=0.031) and separately MI (HR 1.14: 1.04 to 1.26, p=0.008) whilst on Clopidogrel, compared to non-LoF carriers in Cox’s proportional hazards regression models adjusted for age at first Clopidogrel prescription, sex, and the first 10 genetic principal components of ancestry. For this analysis non-LoF carriers constituted those with a ‘normal’ *CYPC219* genotype and those with the *CYP2C19*17* gain-of-function genotype. An in-depth analysis in our companion paper (Supplementary Table 3 in [7]) showed that normal and *\*17* individuals had a near-identical risk of stroke (HR=0.99, p=0.97) and that removing *\*17* individuals had little impact on the analysis estimates other than a loss in precision, since they constitute 22% of the population. For this reason we chose to keep the binary LoF/non-LoF genetic classification for the full TWIST analysis in the next section.

#### 4.1.1 Estimating the GMTE

To estimate the GMTE in this case we modelled the time to stroke using an Aalen additive hazards model, as described in Section 2.4 and *S1 Text*. All models were adjusted for age at recruitment or first Clopidogrel prescription, sex, and the first 10 genetic principal components of ancestry. Fig 7 and Table 5 show the results for this analysis, which reflect the genetically moderated effect of Clopidogrel treatment on the hazard of stroke per year, expressed as a percentage. The GMTE(1) estimate suggests that being a CYP2C19 LoF carrier (*G*=1) increases the risk of stroke by 0.28% (p=0.048) compared to those without the LoF variant (*G*=0). To put this figure in context, if we could reduce the LoF carrier’s risk by this amount then, when multiplied by the 5264 LoF carrier patient years in the data, it would lead to an expected 13.2% reduction in the total number of strokes (or a reduction of 15 strokes from 110 to 95).

**Table 5:** Hazard difference estimates (LoF carriers versus non-carriers) on percentage scale for all single and combined estimates

| Estimator | Estimate<br>(% scale) | S.E | p-value | Combined Estimates |  |  |
| --- | --- | --- | --- | --- | --- | --- |
|  |  |  |  | Q statistic | Q p-value | Combine? |
| CAT | 2.2 | 0.210 | 0.0e+00 |  |  |  |
| GMTE(0) | $-3.9 \times 10^{-3}$ | $7.5 \times 10^{-3}$ | 6.1e-01 | | | |
| GMTE(1) | 0.28 | 0.140 | 4.8e-02 |  |  |  |
| MR | 0.29 | 0.110 | 7.9e-03 |  |  |  |
| RGMTE | 0.33 | 0.160 | 3.7e-02 |  |  |  |
| RGMTE/MR | 0.3 | 0.089 | 7.5e-04 | 0.05 | 8.3e-01 | Yes |
| RGMTE/CAT | 1.0 | 0.130 | 2.0e-15 | 49 | 2.4e-12 | No |
| MR/CAT | 0.68 | 0.096 | 9.1e-13 | 64 | 1.3e-15 | No |
| GMTE(1)/CAT | 0.86 | 0.120 | 1.0e-13 | 56 | 5.9e-14 | No |
| RGMTE/MR/CAT | 0.59 | 0.082 | 6.6e-13 | 68 | 2.1e-15 | No |

**Fig 7:**
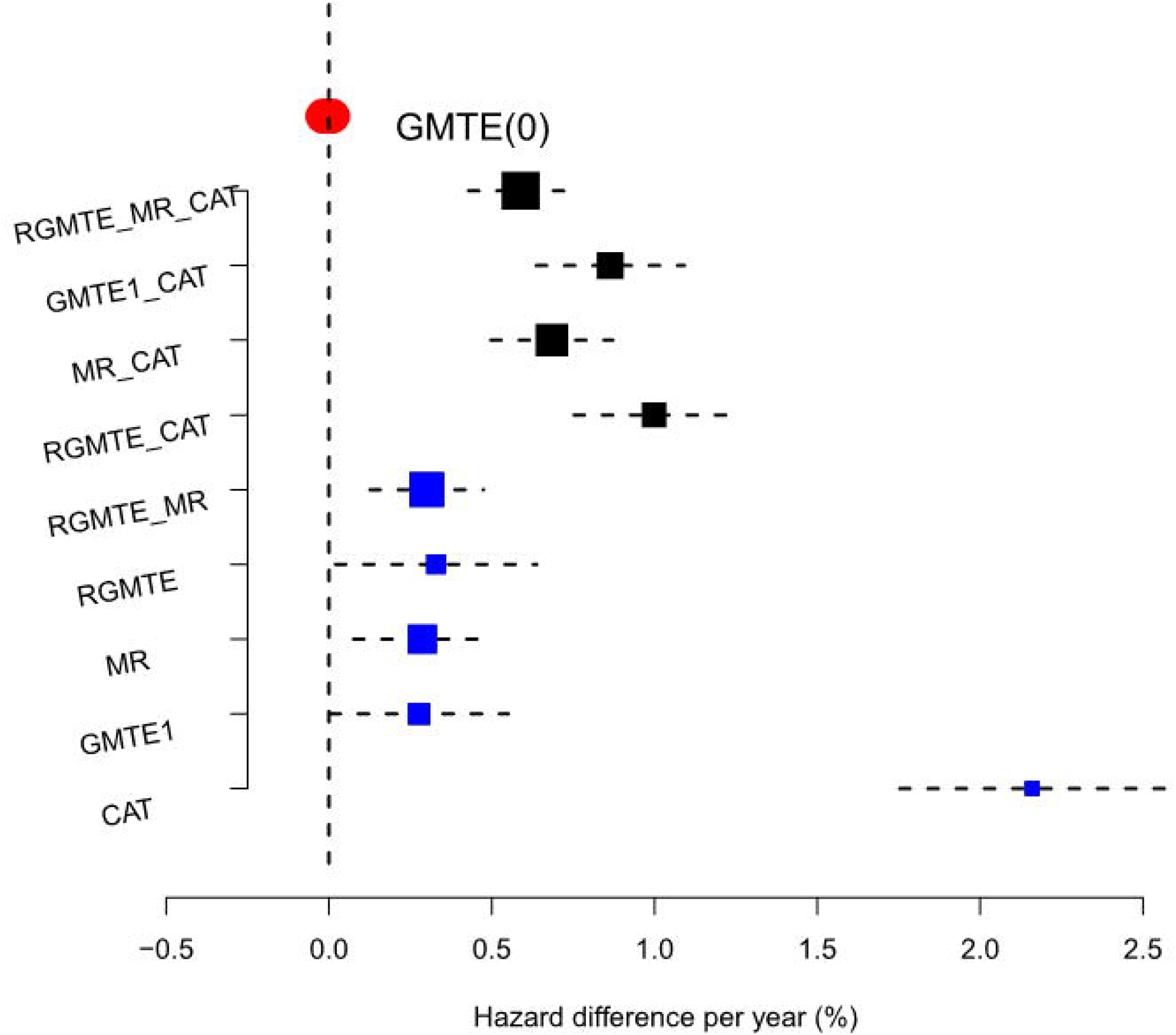
Forest plot of Results for the Clopidogrel data. Blue squares show individual causal estimates as well as combined estimators that pass the heterogeneity test at the 5% level. Black squares show combined estimates that fail the heterogeneity test at the 5% level. Red bar shows the point estimate and confidence interval for the GMTE(0) estimate.

To test for potential bias in the GMTE(1) estimate, we calculate the GMTE(0) estimate in the untreated population. Thankfully, it is close to zero (Hazard diff = -0.0039%, p=0.61), although slightly negative. Taken at face value, this suggest LoF carriers have a slightly reduced risk of stroke through pathways other than Clopidogrel use. Next we calculate the Corrected As Treated (CAT) estimate. As discussed, the validity of this method rests strongly on being able to identify all confounders of Clopidogrel use and stroke. With the data available, it was only possible to adjust for age, sex and genetic principal components and perhaps unsurprisingly, the CAT estimate is an order of magnitude larger (Hazard diff = 2.2%, p ≤ 2 × 10^*−*16^). Consequently, the *Q*_*CAT,GMT E*(1)_ statistic detects large heterogeneity and suggests that the CAT and GMTE(1) estimates should not be combined.

For completeness, we next calculate the RGMTE estimate for the GMTE hazard difference. Since this is itself the difference between the GMTE(1) and GMTE(0) estimates, and given they are of opposite sign, the RMGTE estimate is slightly larger at 0.33% (p=0.037), suggesting 17 strokes could have been avoided. The MR estimate for the GMTE hazard difference is similar at 0.29% (p=0.008). Heterogeneity analysis reveals that the MR and RGMTE estimates are sufficiently similar to combine into a more precise single estimate of the GMTE (*Q*_*MR,RGMT E*_=0.8). The combined estimate is 0.3 (p=7.5 × 10^*−*04^), or that 16 strokes could have been avoided. No other combination of estimates are sufficiently similar to combine.

### 4.2 Statins, *APOE* & CAD

We now apply our framework to estimate the extent to which genetic variation at the *APOE* locus modulates the risk of coronary artery disease (CAD) due to statin treatment using UK Biobank data. Our full data comprises 155,409 unrelated participants of European descent, with primary care data available (updated to March 2017) and up-to-date hospital admission data as of December 2020. Of this sample, we excluded: n=11 participants with missing *APOE* genotypes; n=6,456 non-regular statin users with less than four prescriptions per year or residuals from the linear regression for total statin prescriptions on years of statin treatment greater than 3 or less than -3; n=1,273 non-statin users diagnosed with CAD at baseline (or prior to baseline); n=4,566 participants starting statin after a doctor’s diagnosis of coronary artery disease (CAD) based on the hospital admission records.

Among the included samples (n=143,103), 57,682 (59.5%) were female. Of these, 46,179 (32.3%) were statin users, with a median of 9.4 (inter-quartile range: 6.6 to 13.5) statin prescriptions per year and a median of 5.6 (inter-quartile range: 1.2 to 9.9) years of statin treatment. Several SNPs were associated with LDL cholesterol response to statins based on a genome-wide association study, where the *APOE* e2 defining SNP rs7412 showed a larger LDL cholesterol lowering response to statins compared to e3e3s [16]. APOE genotypes (diplotypes essentially) were determined based on genotypes at rs7412 and rs429358. Inspecting the *APOE* genotype distribution, the majority of participants were classed as *e*_3_*e*_3_ (n=83,813, 58.6%), followed by *e*_3_*e*_4_ (n=33,597, 23.5%), *e*_2_*e*_3_s (n=17,811, 12.4%), *e*_2_*e*_4_s (n=3,616, 2.5%), *e*_4_*e*_4_s (n=3,366, 2.4%), and *e*_2_*e*_2_s (n=900, 0.6%). These groups are mutually exclusive. Summary statistics for statin users and non-statin users are presented in Table 6.

**Table 6:** Baseline covariates, genetic data and incident CAD cases on statin users and non-users in UK Biobank.

|  | Statin Users (n=46,179) | Non-Statin Users (n=96,924) |
| --- | --- | --- |
| Age starting statin (left) |  |  |
| age at recruitment (right) | 60.5± 7.9 | 54.7± 8.0 |
| Sex (=Female) | 20,921 (45.3%) | 57,682 (59.5%) |
| *APOE* genotype |  |  |
| *e*<sub>3</sub>*e*<sub>3</sub> | 26,938 (58.3%) | 56,875 (58.7%) |
| *e*<sub>2</sub>*e*<sub>2</sub> | 258 (0.6%) | 642 (0.7%) |
| *e*<sub>2</sub>*e*<sub>3</sub> | 4,772 (10.3%) | 13,039 (13.5%) |
| *e*<sub>2</sub>*e*<sub>4</sub> | 1,065 (2.3%) | 2,551 (2.6%) |
| *e*<sub>3</sub>*e*<sub>4</sub> | 11,849 (25.7%) | 21,748 (22.4%) |
| *e*<sub>4</sub>*e*<sub>4</sub> | 1,297 (2.8%) | 2,069 (2.1%) |
| Incident CAD (MI or angina) cases | 7,259 (15.7%) | 2,758 (2.8%) |

### 4.3 Results

Using the *e*3*e*3 group as a reference, we fitted Aalen additive hazard models within each mutually exclusive genetic group additionally adjusting for sex, age on statin or age at recruitment, and the top 10 genetic principal components. For brevity, we focus on the results of the e2e3 versus e3e3 and e4e4 versus e3e3 analyses, which are shown in Table 7 and account for approximately 72% of the patient data. Estimates reflect the hazard or risk difference of a CAD event per year, expressed as a percentage. Only results of combined estimates that pass a heterogeneity test at the 5% level are shown. Equivalent estimates for the remaining genetic groups showed no evidence of a non-zero genetically moderated effect. Results for all genetic groups are given in Table 8.

**Table 7:** Hazard difference estimates on the % scale for all single and valid combined estimates for the e2e3 and e4e4 genotype groups

| Estimator | Estimate | S.E | p-value | Expected Avoided<br>CAD events in<br>e3e3 group (95% CI) |
| --- | --- | --- | --- | --- |
| **e4e4 versus e3e3** |  |  |  |  |
| CAT | 3.9000 | 0.082 | 0.000 | - |
| GMTE1 | -0.0310 | 0.015 | 0.043 | -85 (-168, -3) |
| MR | 0.0069 | 0.017 | 0.690 | 19 (-76,115) |
| RGMTE | -0.0370 | 0.018 | 0.046 | -103 (-204,-2) |
| GMTE0 | 0.0110 | 0.0063 | 0.073 | - |
| RGMTE/MR | -0.0140 | 0.013 | 0.280 | -39 (-108,31) |
| **e2e3 versus e3e3** |  |  |  |  |
| CAT | 1.2000 | 0.0240 | 0.0e+00 | - |
| GMTE1 | -0.0100 | 0.0087 | 2.4e-01 | -28 (-75,19) |
| MR | -0.0460 | 0.0110 | 3.4e-05 | -128 (-189,-67) |
| RGMTE | -0.0055 | 0.0098 | 5.7e-01 | -15 (-69,38) |
| GMTE0 | -0.00014 | 0.0025 | 9.5e-01 | - |

**Table 8:**
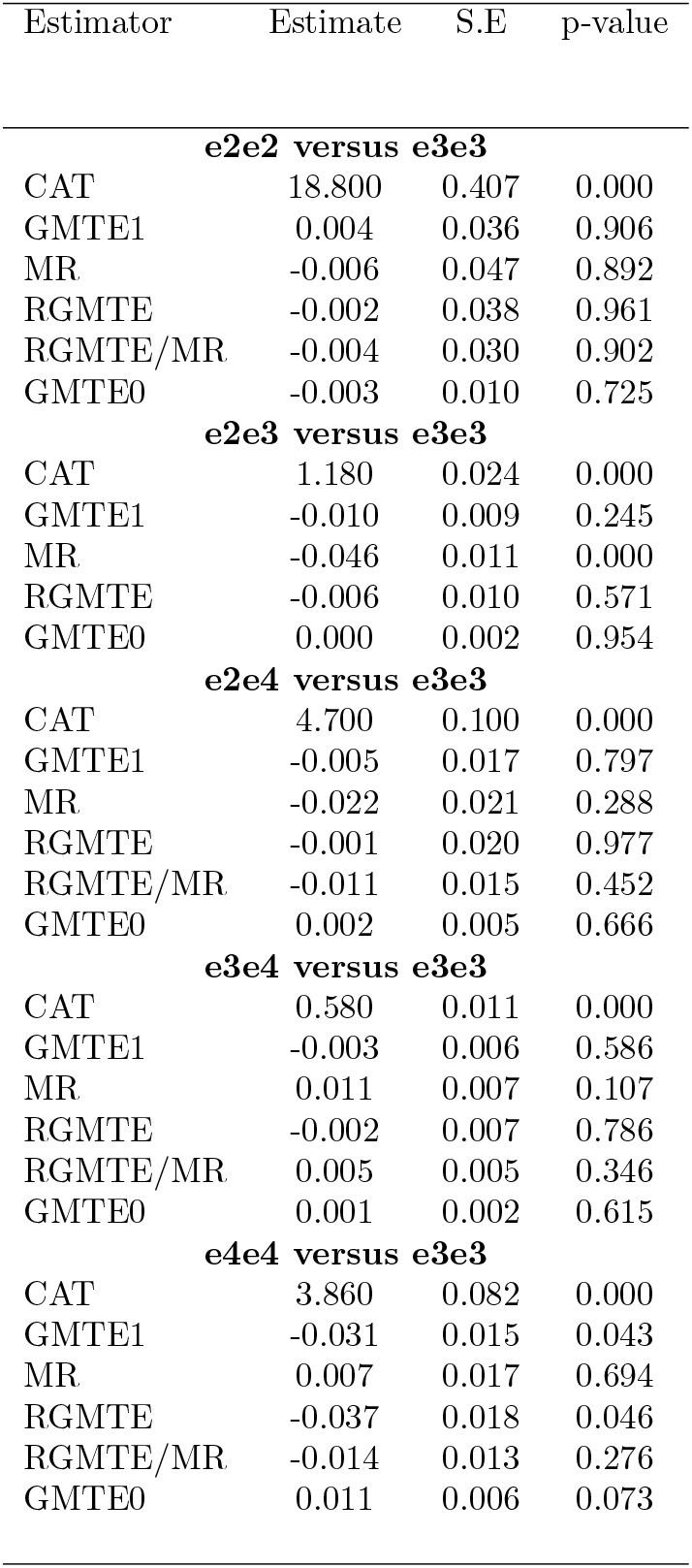
Hazard difference estimates on the % scale for all single and valid combined estimates

#### 4.3.1 *e*_4_*e*_4_ versus *e*_3_*e*_3_

Inspecting the *e*_4_*e*_4_ genetic subgroup first, the GMTE(1) estimate suggests that the risk of CAD could be reduced by 0.031% per year if *e*_3_*e*_3_ patients experienced the same treatment effect as *e*_4_*e*_4_ patients (p=0.043). This estimate is valid if the e4e4 genotype only affects the risk of CAD through modulating the effectiveness of statins (i.e. assumptions PG1-PG3 hold). In order to probe this we calculate the equivalent GMTE(0) estimate in non-statin users. The *e*_4_*e*_4_ group is now seen to have a 0.011% larger risk of CAD than *e*_3_*e*_3_ (p=0.07), which suggests that PG1-PG3 violation is possible. Furthermore, Table 6 shows clear differences in the allele frequencies between treatment groups. Since the RMGTE(0) and RGMTE(1) estimates are of opposite signs, the RGMTE estimate, which is robust to PG2-PG3 violation, infers the risk difference between e4e4 and e3e3’s is larger at -0.037% per year (p=0.046). The MR estimate of the GMTE is also positive (0.0069%), but very close to zero (p=0.69). This is, however, sufficiently similar to the RGMTE estimate for it to be combined with the MR estimate, and the combined value suggests a hazard difference of -0.014% per year (p=0.28). The CAT estimate for the hazard difference in these data is a 3.9% increase per year. Its magnitude is so large compared to the other estimates that we can infer adjustment for age, sex and genetic PCs is again not sufficient to remove confounding by indication. Consequently, no other estimate is sufficiently similar in order to combine with the CAT estimate, as shown in Fig 8. In the final column of Table 7 we translate the hazard difference estimate per year implied by the GMTE1, MR, RGMTE and combined RMGTE/MR estimate to give an expected number of CAD events that could be avoided if all 26,938 e3e3 statin user patients could receive the same benefit as the e4e4 patients, by multiplying the per-year risk reduction over the relevant 278,409 patients-years in the data. Using the RGMTE estimate for this risk reduction gives a figure of 103. The GMTE1 and combined RGMTE/MR estimates imply more modest reductions of 85 and 39 CAD events respectively.

**Fig 8:**
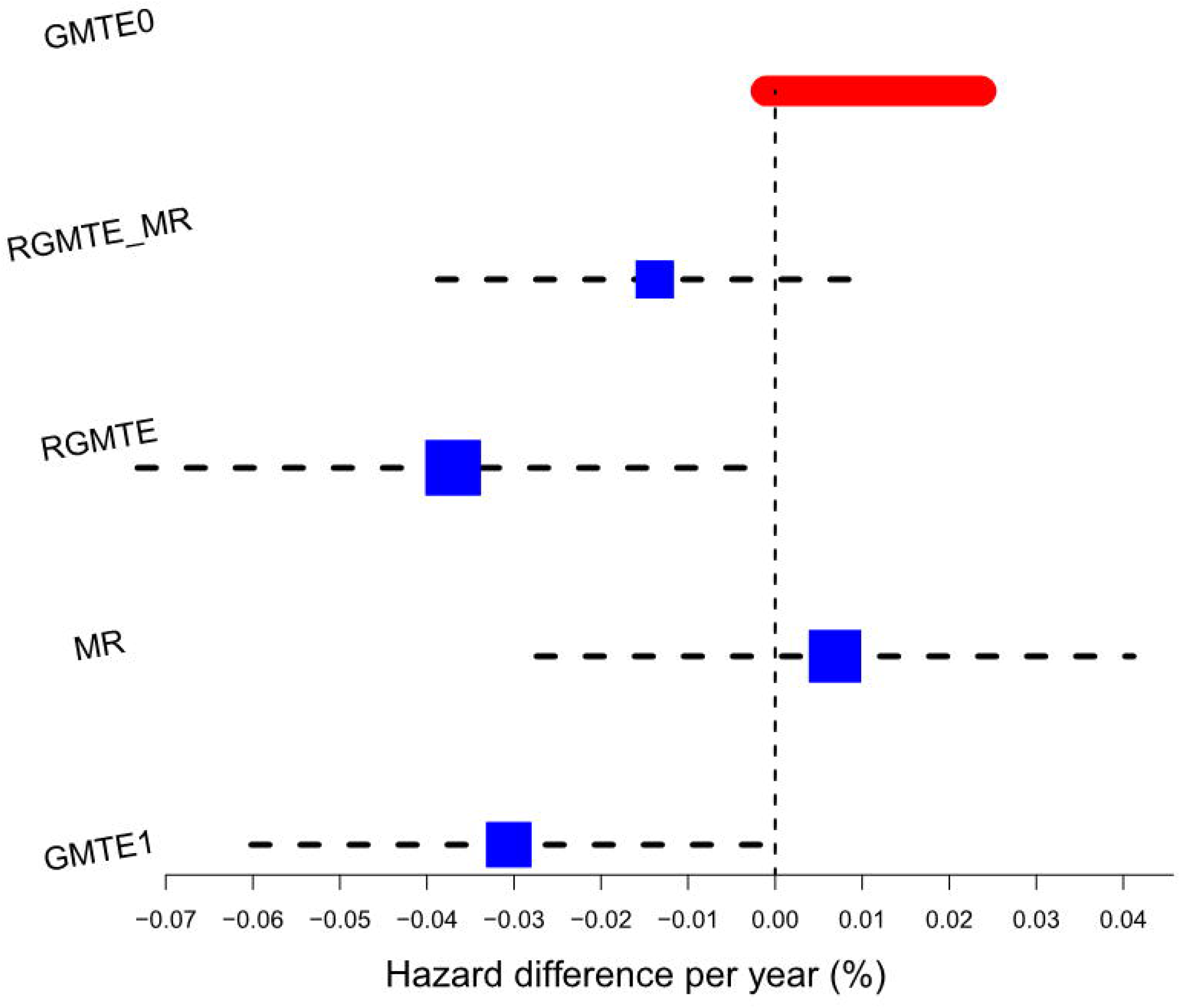
Hazard difference estimates for the *e*_4_*e*_4_ versus *e*_3_*e*_3_ analyses. Color coding the same as for Figure 7.

#### 4.3.2 *e*_2_*e*_3_ versus *e*_3_*e*_3_

Turning our attention to the e2e3 subgroup in Table 7 and Fig 9, we again see a large, non-credible CAT hazard difference estimate for the GMTE of a 1.2% per year between e2e3 and e3e3 groups. The GMTE(1), GMTE(0) and RGMTE estimates for the GMTE are all close to zero and non-significant at the 5% level. In contrast, the MR estimate for the GMTE suggests that e2e3’s have a 0.046% reduced risk of CAD per year (p=3 × 10^*−*5^). Using this estimate, the expected number of CAD events that could be avoided if all 26,938 e3e3 patients could receive the same benefit as the e2e3 patients is 128. This is valid if we believe that assumptions PG2-PG3 hold, but either PG1 or the Homogeneity assumption are violated. The GMTE1 and RGMTE estimates imply more modest reductions of 28 and 15 CAD events respectively. In this example, no two single uncorrelated estimates are sufficiently similar in order to combine.

**Fig 9:** Hazard difference estimates for the *e*_2_*e*_3_ versus *e*_3_*e*_3_ (right) analyses. Color coding the same as for Figure 7.

## 5 Discussion

In this paper we propose the general TWIST framework for estimating the genetically moderated treatment effect that combines several distinct but complementary causal inference techniques. We propose a rudimentary decision framework for choosing when to combine approaches based on heterogeneity statistics. In practice, expert knowledge and prior evidence should also be leveraged to decide whether the particular assumptions of the causal estimation strategy are likely to be met, in order to put more or less weight on their findings. For example, if a variant is known to be associated with the outcome through another mechanistic pathway, then the PG3 assumption required for the GMTE(1) and MR estimates is likely violated, and the RGMTE estimate should be favored. Or, if it is known that those with the metabolically unfavourable genotype (*G*=0) still benefit from treatment, then the homogeneity assumption is likely violated. This would then rule out the CAT estimate completely and one would need to be sure the PG1 assumption was satisfied when using the MR estimate.

In *S1 Code* we provide R code for fitting the TWIST framework for continuous, binary and time-to-event data as well as code used in the simulation study. Work is ongoing to produce a single R package to apply TWIST and visualise its results. Our inverse variance meta-analysis procedure for combining estimates is very simple, and simulations showed that exhibited good statistical properties even when small correlations between constituent estimates were present. As future work, we plan to develop a more sophisticated procedure to explicitly account for this correlation within TWIST based on a Mahalanobis distance statistic, and to further develop the framework in several directions to address current limitations, some of which are now described.

### 5.1 Limitations and further work

We chose to illustrate the utility of the TWIST framework for combining similar estimates by demonstrating that it can increase precision. An alternative strategy would be to use multiple estimates to improve the robustness of any inference due to possible violations of variety of assumptions. For example, given a prior null hypothesis about the specific value of the GMTE, we would not reject the hypothesis it if was not rejected by any of the individual analysis. On the other hand, we could reject a proposed value of the estimand with increased confidence if it is rejected by multiple independent analyses that depend on assumptions that do not completely overlap. In future work we plan to develop a rigorous sequential testing procedure for TWIST that can control family wise error or false discovery rates. We thank reviewer 1 and 4 for these helpful suggestions.

The TWIST framework offers a means to combine statistically uncorrelated estimates that rely on overlapping sets of assumptions. If two estimates are similar enough to warrant combining into a single estimate, one hopes that this represents a more precise estimate of the true GMTE. However, there is always the possibility that both estimates are instead systematically biased in the same direction when there is a degree of overlap in their identifying assumptions and these assumptions are violated. In this case, combining them could give a more precise estimate of the wrong answer. Although we saw little evidence of this in simulation scenarios 4-6, further research is needed to understand the extent of this issue more clearly. We thank reviewer 4 for raising this important point.

In our analysis of the statin data, we estimated the GMTE in several mutually exclusive genetic groups, which resulted in an inevitable loss of precision. Efficiency could potentially be regained by collapsing genetic subsets together if they give similar estimates, or by making a linearity assumption about the magnitude of effect across genotypic groups (e.g. between *e*_3_*e*_3_, *e*_3_*e*_4_ and *e*_4_*e*_4_). This would not be defensible if the genetic groups were ordered with respect to the magnitude of their causal estimate, but would be defensible if genetic groups could be ordered by their effect on increasing drug metabolism. In the case of 3 genetic groups, *G*_*i*_ and 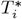 could take a value in {0,1,2}. This would enable the data to be pooled in order to target a combined estimand

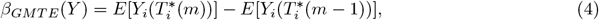

for all *m* in {1, 2}. If such a model were correct, it opens up the possibility of making the analysis even more robust to violations of the PG assumptions, because an additional causal parameter could be jointly estimated alongside the GMTE to reflect, for example the direct effect of *G* on *Y*. This is an important avenue for further research.

Although the CAT estimate can, in principle consistently estimate the GMTE estimand, it relies heavily on the NUC assumption. In both applied analyses we were not able to sufficiently control for confounding by indication to deliver an estimate close to any other GMTE estimate, due to a lack of relevant covariate data. In future work we plan to revisit both analyses after collecting a much larger set of relevant information. More-sophisticated approaches such as Propensity Scores, matching methods and inverse probability weighting may then offer some utility [17]. So too may methods for multi-variable Mendelian randomization, where instead of directly adjusting for confounders of treatment and outcome, we instead adjust for their genetically predicted value. This latter approach could be more robust to collider bias [11].

The TWIST framework has parallels with the general theory of ‘Evidence Factors’ [18] for combining two or more observational associations estimates gleaned from the same data, which are susceptible to different biases. As far as we are aware, this approach has not been applied within the context of pharmacogenetics before, but a more detailed investigation of the connection between TWIST and Evidence Factors is an interesting topic for further research.

## Supporting information

technical details

## Data Availability

The data used in this paper comes from UK Biobank, which is publicly available to all bona fide researchers worldwide following a formal application

## Figure Legends

S1 Text: Document containing the important technical details on the TWIST framework, including consistency proofs for the linear case, and the implementation of TWIST with binary and time-to-event data.

S1 Code: Zip file containing code to implement the TWIST framework with continous, binary and time-to-event data.

