## Supplementary material for "The Triangulation WIthin A STudy (TWIST) framework for causal inference within Pharmacogenetic research": technical details


### Consistency of the CAT estimate

The CAT(Y) estimand is equal to

$$\beta_{CAT}(Y) = \frac{E[Y|T=1] - E[Y|T=0]}{E[G|T=1]},$$

Under model (2) the numerator of the estimand equals

$$\begin{aligned}
& \beta_0 + (\beta_1 - \beta_0)E[G|T=1] + \gamma_{YG}E[G|T=1] + \gamma_{YU}E[U|T=1] - (\gamma_{YG}E[G|T=0] + \gamma_{YU}E[U|T=0]) \\
= \quad & \beta_0 + \beta_{GMTE}E[T|G=1] + \gamma_{YG}\left\{E[G|T=1] - E[G|T=0]\right\} + \gamma_{YU}\left\{E[U|T=1] - E[U|T=0]\right\}
\end{aligned}$$

Dividing through by $E[G|T=1]$ we see that the full estimand equals

$$\beta_{CAT}(Y) = \beta_0 + \beta_{GMTE} + \gamma_{YG}\left(1 - \frac{E[G|T=0]}{E[G|T=1]}\right) + \gamma_{YU}\beta_{CAT}(U)$$

The CAT estimand is therefore equivalent to $\beta_{GMTE}$ if the Homogeneity and the NUC assumptions hold, and either PG1 or PG3 hold.

### Consistency of the GMTE(1) estimate

The GMTE(1) estimand under model (2) is equal to

$$\begin{aligned}
E[Y|T=1,G=1] - E[Y|T=1,G=0] \quad &= \quad \gamma_{Y0} + \beta_0 + (\beta_1 - \beta_0) + \gamma_{YG} + \gamma_{YU}E[U|T=1,G=1] \\
&- \quad (\gamma_{Y0} + \beta_0 + \gamma_{YU}E[U|T=1,G=0]) \\
&= \quad (\beta_1 - \beta_0) + \gamma_{YG} + \gamma_{YU}\left\{E[U|T=1,G=1] - E[U|T=1,G=0]\right\} \\
&= \quad \beta_{GMTE(1)} + \gamma_{YG} + \gamma_{YU}\beta_{GMTE(1)}(U)
\end{aligned}$$

For this estimand to equal $\beta_{GMTE}$ we require assumption PG3 ($\gamma_{YG}$=0) and that $G \perp\!\!\!\perp U|T=1$. Since PG1 and PG2 imply that $G \perp\!\!\!\perp (U,T)$, the GMTE(1) estimate is consistent for the GMTE under PG1-PG3.

Under model (2) the GMTE(0) estimand is equal to

$$\begin{aligned}\hat{\beta}_{GMTE}(0) = E[Y|T=0,G=1] - E[Y|T=0,G=0] &= \gamma_{Y0} + \gamma_{YG} + \gamma_{YU} E[U|T=0,G=1] \\ &- (\gamma_{Y0} + \gamma_{YU} E[U|T=0,G=0]) \\ &= \gamma_{YG} + \gamma_{YU} \{E[U|T=0,G=1] - E[U|T=0,G=0]\} \\ &= \gamma_{YG} + \gamma_{YU} \beta_{GMTE(0)}(U)\end{aligned}$$

This estimand will equal zero when PG3 ($\gamma_{YG}$=0) and $G \perp\!\!\!\perp U|T=0$ hold. Using the same logic as above the GMTE(0) estimate is a consistent estimate for 0 under PG1-PG3.

### Consistency of the RGMTE estimate

Under model (2) we have that RGMTE(Y) estimand is equal to

$$\begin{aligned} & E[Y|T=1,G=1] - E[Y|T=1,G=0] - (E[Y|T=0,G=1] - E[Y|T=0,G=0]) \\ = \; & (\beta_1 - \beta_0) + \gamma_{YG} + \gamma_{YU} \{E[U|T=1,G=1] - E[U|T=1,G=0]\} \\ - \; & \gamma_{YG} + \gamma_{YU} \{E[U|T=0,G=1] - E[U|T=0,G=0]\} \\ = \; & \beta_{GMTE} + \gamma_{YU} (\{E[U|T=1,G=1] - E[U|T=1,G=0]\} - \{E[U|T=0,G=1] - E[U|T=0,G=0]\}) \\ = \; & \beta_{GMTE} + \gamma_{YU} \beta_{RGMTE}(U)\end{aligned}$$

The RGMTE(Y) estimand is therefore equivalent to $\beta_{GMTE}$ if the RGMTE(U) estimand is zero. This will be zero whenever the strength of association between $U$ and $G$ is independent of $T$ or that

$$Cov(U,G)|T = Cov(U,G)$$

Note that this second condition will be satisfied if either PG1 holds or NUC holds. If both assumptions are violated then conditioning on treatment induces a collider bias which alters the strength of association between $U$ and $G$.

### Consistency of the MR estimate

Under model (2) the MR estimand is equal to:

$$\beta_{MR}(Y) = \frac{E[Y|G=1] - E[Y|G=0]}{E[T^*|G=1] - E[T^*|G=0]}$$

We first note that the denominator of the MR estimand simplifies to $E[T=1|G=1]$ since $E[T^*|G=1] = E[TG|G=1] = E[T|G=1]$ and that $E[T^*|G=0] = E[TG|G=0] = 0$. We can then write the estimand as

$$\begin{aligned}\beta_{MR}(Y) = \frac{E[Y|G=1] - E[Y|G=0]}{E[T|G=1]} &= (\beta_1 - \beta_0) \\ &+ \frac{\beta_0(E[T|G=1] - E[T|G=0])}{E[T|G=1]} \\ &+ \frac{\gamma_{YU}(E[U|G=1] - E[U|G=0])}{E[T|G=1]} \\ &+ \frac{\gamma_{YG}}{E[T|G=1]} \\ &= \beta_{GMTE} \\ &+ \beta_0 \left(1 - \frac{E[T|G=0]}{E[T|G=1]}\right) + \gamma_{YU}\beta_{MR}(U) + \frac{\gamma_{YG}}{E[T|G=1]}\end{aligned}$$

From this we can see that the consistency of the MR estimate relies on assumption PG3 being satisfied in order for the third bias term to disappear, but there are two possible ways for the first and second bias terms to disappear. For the first bias term, either assumption PG1 is satisfied, so that $E[T|G=1]$-$E[T|G=0] = 0$, or the Homogeneity assumption is satisfied, so that $\beta_0 = 0$. For the second bias term to disappear we either require the NUC assumption or PG2.

### Estimating the GMTE with binary and time-to-event data

When the outcome is continuous, the TWIST analysis approaches can be implemented using linear regression to estimate the GMTE as a mean difference. With a binary outcome, we recommend estimating risk differences directly using either a linear probability model, or using a logistic regression model to furnish estimates on the risk difference scale. In the latter case, a risk difference can be constructed by calculating the mean difference between predicted probabilities under manipulation of the appropriate variable between levels 1 and 0, as suggested by Gelman and Hill [1]. For example, in the case of the GMTE(1) estimator

$$\hat{\beta}_{GMTE(1)}(Y) = \frac{1}{n}\sum_{i=1}^{n} \hat{\pi}(Y)(T^* = 1, T = 1, Z = z_i) - \hat{\pi}(Y)(T^* = 0, T = 1, Z = z_i)$$

where $\hat{\pi}(Y)(T^* = t_i^*, T = j, Z = z_i)$ is the estimated fitted value from a logistic regression of $Y$ on $T$,$T^*$ and $Z$ at treatment level $j$. For time-to-event data, we recommend analysing the data using an Aalen additive hazard model, as suggested by Tchetgen et. al. [2]. Specifically, one would assume that model (**??**) holds on the additive hazard scale, so that at time $t_y$ the hazard:

$$h(t_y|T, U, G, Z) = \gamma_{Y0} + \beta_1 TG + \beta_0 T(1 - G) + \gamma_{YG}G + \gamma_{YU}U + \gamma_{YZ}Z \quad (1)$$

### Which estimates are uncorrelated?

Let $\bar{Y}_{ij} = \hat{E}[Y|T = i, G = j]$, $\text{Var}(\bar{Y}_{ij}) = \sigma^2$, $\pi_{ij} = \hat{E}[T = i|G = j]$ and $\tau_{ij} = \hat{E}[G = j|T = i]$. We now write the CAT, GMTE(1), RGMTE and MR estimates as

$$\begin{aligned}
\hat{\beta}_{CAT(Y)} &= \frac{\bar{Y}_{10}\tau_{10} + \bar{Y}_{11}\tau_{11} - (\bar{Y}_{00}\tau_{00} + \bar{Y}_{01}\tau_{01})}{\tau_{11}} \\
\hat{\beta}_{GMTE(1)}(Y) &= \bar{Y}_{11} - \bar{Y}_{10} \\
\hat{\beta}_{MR(Y)} &= \frac{\bar{Y}_{11}\pi_{11} + \bar{Y}_{01}\pi_{01} - (\bar{Y}_{10}\pi_{10} + \bar{Y}_{00}\pi_{00})}{\pi_{11}} \\
\hat{\beta}_{RGMTE}(Y) &= \bar{Y}_{11} - \bar{Y}_{10} - (\bar{Y}_{01} - \bar{Y}_{00})
\end{aligned}$$

Ignoring the denominator of $\hat{\beta}_{MR}$, the covariance between its numerator and $\hat{\beta}_{RGMTE}$ can be shown to equal zero. Let $n_{ij}$ equal the number of subjects with variables $T = i$ and $G = j$ in the sample and let $n_{Gj}$ equal the number of subjects with variable $G = j$ in the sample. It then follows that:

$$\begin{aligned}
\pi_{11}\text{Cov}(\hat{\beta}_{MR}(Y), \hat{\beta}_{RGMTE}(Y)) &= \pi_{11}\text{Var}(\bar{Y}_{11}) - \pi_{10}\text{Var}(\bar{Y}_{10}) - \pi_{01}\text{Var}(\bar{Y}_{01}) + \pi_{00}\text{Var}(\bar{Y}_{00}) \\
&= \pi_{11}\frac{\sigma^2}{n_{11}} + \pi_{10}\frac{\sigma^2}{n_{10}} - \pi_{01}\frac{\sigma^2}{n_{01}} - \pi_{00}\frac{\sigma^2}{n_{00}} \\
&= \frac{\sigma^2}{n_{G1}} + \frac{\sigma^2}{n_{G1}} - \frac{\sigma^2}{n_{G0}} - \frac{\sigma^2}{n_{G0}} \\
&= 0
\end{aligned}$$

Using similar arguments it is easy to show that

$$\begin{aligned}
\tau_{11}\text{Cov}(\hat{\beta}_{CAT}(Y), \hat{\beta}_{RGMTE}(Y)) &= 0 \\
\tau_{11}\pi_{11}\text{Cov}(\hat{\beta}_{CAT}(Y), \hat{\beta}_{MR}(Y)) &= 0 \text{ if } G \perp\!\!\!\perp T, \\
\text{Cov}(\hat{\beta}_{CAT}(Y), \hat{\beta}_{GMTE(1)}(Y)) &= 0
\end{aligned}$$

In cases where $G \perp\!\!\!\perp T$ does not hold, but $G$ predicts a relatively small amount of variation in $T$, the correlation between the CAT and MR estimate will be non-zero but negligible. This is a reasonable assumption in almost all pharmacogenetic contexts. This means that there are effectively only two pairs of correlated estimates: (GMTE(1), RGMTE), and (GMTE(1), MR).

#### Combining uncorrelated estimates

In order to decide whether two uncorrelated estimates can be combined, we propose the use of a simple heterogeneity statistic. This procedure is illustrated in Fig **??** (left) taking the GMTE(1) and CAT estimates as an example. Using each estimate we calculate their inverse variance weighted average and from this the heterogeneity statistic, $Q_{GMTE(1),CAT}$.

$$Q_{GMTE(1),CAT} = \sum_e w_e(\hat{\beta}_e(Y) - \hat{\beta}_c(Y))^2, \text{ where } \hat{\beta}_c(Y) = \frac{\sum_e w_e \hat{\beta}_e}{\sum_e w_e}, \quad e = \{GMTE(1), CAT\},$$

and where $w_e$ is the inverse variance of estimate $\hat{\beta}_e$. If this statistic is less than 1-$\alpha$ quantile of a $\chi^2_d$ (where $\alpha$ is the pre-specified significance threshold and $d$ = Card($e$)-1 = 1) then we judge the GMTE(1) and CAT estimates to be sufficiently similar to combine into a single estimate $\hat{\beta}_{GMTE(1),CAT}(Y)$. If $Q_{GMTE(1),CAT}$ is greater than 1-$\alpha$ threshold then the two estimates should be left separate.

#### When does $\hat{\beta}_{RGMTE,MR}(Y)$=$\hat{\beta}_{GMTE(1)}(Y)$?

Using the notation from the previous appendix, we can write the MR estimand as

$$\begin{aligned}
\frac{E[Y|G=1] - E[Y|G=0]}{E[T|G=1]} &= \frac{\bar{Y}_{01}\pi_{01} + \bar{Y}_{11}\pi_{11} - (\bar{Y}_{00}\pi_{00} + \bar{Y}_{10}\pi_{10})}{\pi_{11}} \\
&= \frac{\bar{Y}_{11}\pi_{11} - \bar{Y}_{10}\pi_{10} + (\bar{Y}_{01}\pi_{01} + \bar{Y}_{00}\pi_{00})}{\pi_{11}}
\end{aligned}$$

Under the assumption that $G \perp\!\!\!\perp T$ we have that $\pi_{10} = \pi_{11} = Pr(T=1) = \pi_1$, $\pi_{00} = \pi_{01} = Pr(T=0)$ $= \pi_0$ and the MR estimand equals

$$\beta_{GMTE(1)}(Y) + \beta_{GMTE(0)}(Y)\frac{\pi_1}{\pi_0}$$

The RGMTE estimand is $\beta_{GMTE(1)}(Y) - \beta_{GMTE(0)}(Y)$. Therefore, when additionally $Pr(T=1) = Pr(T=0)$, both the RGMTE and MR estimates will have equal precision and the GMTE(1) estimate will be the average of the RGMTE estimand and the MR estimand. In general, an inverse variance weighted average of the RGMTE and MR estimates is likely to be a close approximation to the GMTE(1) estimate.
