## Supplementary material for "The Triangulation WIthin A STudy (TWIST) framework for causal inference within Pharmacogenetic research": code: BinaryDataCode.R

```
library(margins)

Combine = function(Ests=Ests,SEs=SEs,alpha=0.05){
w           = 1/SEs^2
ws          = w/sum(w)
E           = sum(ws*Ests)
SE          = sqrt(sum((ws*SEs)^2))

E2  = (Ests-E)^2
Q   = sum(w*E2)
df  = length(Ests)-1
Qp  = 1-pchisq(Q,df)
Dec = 0 ; if(Qp>=alpha){Dec=1}


P.Est          = 2*(1-pnorm(abs(E/SE)))
R              = c(E,SE,P.Est,Q,Qp,Dec)
ResultsX       = matrix(R,nrow=1)
return(ResultsX)
}


##############
# Data input #
##############

# Y = outcome (binary)
# T = treatment (binary)
# G = genotype (binary)
# Z = covariates 
# Link = c("logit","probit","identity")


GMTE_binary = function(Y,T,G,Z,Link="logit"){


Tstar = T*G

# MR

Tshat         = glm(Tstar~G+Z,family=binomial)$fitted
MRfit         = glm(Y~Tshat+Z,family=binomial(link=Link))
MarMR         = summary(margins(MRfit))
MR            = MarMR[1,2]
sMR           = MarMR[1,3]

# Corrected-As treated (CAT)

Tcat   = T*mean(G[T==1])
CATfit = glm(Y~Tcat+Z,family=binomial(link=Link))
MarCAT         = summary(margins(CATfit))
CAT            = MarCAT[1,2]
sCAT           = MarCAT[1,3]

# GMTE(1)

GMTE1fit   = glm(Y~T+Tstar+Z,family=binomial(link=Link))
MarGMTE1   = summary(margins(GMTE1fit))
GMTE1      = MarGMTE1[2,2]
sGMTE1     = MarGMTE1[2,3]


# GMTE(0)

tt        =  (1-T)
ts        =  tt*G
GMTE0fit  = glm(Y~tt+ts+Z,family=binomial(link=Link))
MarGMTE0  = summary(margins(GMTE0fit))
GMTE0     = MarGMTE0[1,2]
sGMTE0    = MarGMTE0[1,3]


# RGMTE

RGMTEfit  = glm(Y~T+Tstar+Tshat+Z,family=binomial(link=Link)) 
MarRGMTE  = summary(margins(RGMTEfit))
RGMTE     = MarRGMTE[3,2]
sRGMTE    = MarRGMTE[3,3]

#######################
# Combined methods    #
#######################

Ests         = c(MR,RGMTE); SEs = c(sMR,sRGMTE)
RGMTE_MR     = Combine(Ests,SEs)
Ests         = c(CAT,RGMTE); SEs = c(sCAT,sRGMTE)
RGMTE_CAT    = Combine(Ests,SEs)
Ests         = c(CAT,MR); SEs = c(sCAT,sMR)
MR_CAT       = Combine(Ests,SEs)
Ests         = c(CAT,GMTE1); SEs = c(sCAT,sGMTE1)
GMTE1_CAT    = Combine(Ests,SEs)
Ests         = c(MR,RGMTE,CAT); SEs = c(sMR,sRGMTE,sCAT)
RGMTE_MR_CAT = Combine(Ests,SEs)


FullCombined     = matrix(nrow=5,ncol=6)
FullCombined[1,] = RGMTE_MR
FullCombined[2,] = RGMTE_CAT
FullCombined[3,] = MR_CAT
FullCombined[4,] = GMTE1_CAT
FullCombined[5,] = RGMTE_MR_CAT

colnames(FullCombined) =c("Est","SE","P.Est","Qstat","Qp","Combine?")
rownames(FullCombined) =c("RGMTE_MR","RGMTE_CAT","MR_CAT","GMTE1_CAT","RGMTE_MR_CAT")


return(list(CAT=MarCAT,GMTE1=MarGMTE1,GMTE0=MarGMTE0,MR=MarMR,RGMTE=MarRGMTE,FullCombined=FullCombined))

}

GMTE_binary(Y,T,G,Z,Link="logit")

GMTE_binary(Y,T,G,Z,Link="identity")
```
