## Supplementary material for "The Triangulation WIthin A STudy (TWIST) framework for causal inference within Pharmacogenetic research": code: ContinuousDataCode.R

```
Combine = function(Ests=Ests,SEs=SEs,alpha=0.05){
w           = 1/SEs^2
ws          = w/sum(w)
E           = sum(ws*Ests)
SE          = sqrt(sum((ws*SEs)^2))

E2  = (Ests-E)^2
Q   = sum(w*E2)
df  = length(Ests)-1
Qp  = 1-pchisq(Q,df)
Dec = 0 ; if(Qp>=alpha){Dec=1}

P.Est          = 2*(1-pnorm(abs(E/SE)))

R        = c(E,SE,P.Est,Q,Qp,Dec)
ResultsX = matrix(R,nrow=1)
return(ResultsX)
}


# GMTE_cont requires

# binary genetic variable G
# binary treatment variable T
# Continuous Outcome variable Y
# a matrix Z of covariates


GMTE_cont = function(Y,T,G,Z){

Tstar = T*G

# MR

Tshat = lm(Tstar~G+Z)$fitted
MRfit = summary(lm(Y~Tshat+Z))
MR    = MRfit$coef[2,1]
sMR   = MRfit$coef[2,2]

# Corrected-As treated (CAT)

Tcat   = T*mean(G[T==1])
CATfit = summary(lm(Y~Tcat+Z))
CAT    = CATfit$coef[2,1]
sCAT   = CATfit$coef[2,2]

# GMTE(1)

GMTE1fit  = summary(lm(Y~T+Tstar+Z))
GMTE1     = GMTE1fit$coef[3,1]
sGMTE1    = GMTE1fit$coef[3,2]

# GMTE(0)

tt        =  (1-T)
ts        =  tt*G
GMTE0fit  = summary(lm(Y~tt+ts+Z))
GMTE0     = GMTE0fit$coef[3,1]
sGMTE0    = GMTE0fit$coef[3,2]


# RGMTE

RGMTEfit  = summary(lm(Y~T+Tstar+Tshat+Z)) 
RGMTE     = RGMTEfit$coef[3,1]
sRGMTE    = RGMTEfit$coef[3,2]


#######################
# Combined methods    #
#######################

Ests         = c(MR,RGMTE); SEs = c(sMR,sRGMTE)
RGMTE_MR     = Combine(Ests,SEs)
Ests         = c(CAT,RGMTE); SEs = c(sCAT,sRGMTE)
RGMTE_CAT    = Combine(Ests,SEs)
Ests         = c(CAT,MR); SEs = c(sCAT,sMR)
MR_CAT       = Combine(Ests,SEs)
Ests         = c(CAT,GMTE1); SEs = c(sCAT,sGMTE1)
GMTE1_CAT    = Combine(Ests,SEs)
Ests         = c(MR,RGMTE,CAT); SEs = c(sMR,sRGMTE,sCAT)
RGMTE_MR_CAT = Combine(Ests,SEs)

FullCombined     = matrix(nrow=5,ncol=6)
FullCombined[1,] = RGMTE_MR
FullCombined[2,] = RGMTE_CAT
FullCombined[3,] = MR_CAT
FullCombined[4,] = GMTE1_CAT
FullCombined[5,] = RGMTE_MR_CAT

colnames(FullCombined) =c("Est","SE","P.Est","Qstat","Qp","Combine?")
rownames(FullCombined) =c("RGMTE_MR","RGMTE_CAT","MR_CAT","GMTE1_CAT","RGMTE_MR_CAT")

return(list(MRfit=MRfit,CATfit=CATfit,GMTE1fit=GMTE1fit,
       GMTE0fit=GMTE0fit,RGMTEfit=RGMTEfit,FullCombined=FullCombined))

}
```
