## Supplementary material for "The Triangulation WIthin A STudy (TWIST) framework for causal inference within Pharmacogenetic research": code: SimulationStudyCode.R

```
rm(list=ls(all=TRUE))

plotfile = c("C:\\Users\\jb1203\\Google Drive\\AExeter\\MR\\Clopidogrel\\Sim1.pdf",
         "C:\\Users\\jb1203\\Google Drive\\AExeter\\MR\\Clopidogrel\\Sim2.pdf",
         "C:\\Users\\jb1203\\Google Drive\\AExeter\\MR\\Clopidogrel\\Sim3.pdf",
         "C:\\Users\\jb1203\\Google Drive\\AExeter\\MR\\Clopidogrel\\Sim4.pdf",
         "C:\\Users\\jb1203\\Google Drive\\AExeter\\MR\\Clopidogrel\\Sim5.pdf",
         "C:\\Users\\jb1203\\Google Drive\\AExeter\\MR\\Clopidogrel\\Sim6.pdf",
         "C:\\Users\\jb1203\\Google Drive\\AExeter\\MR\\Clopidogrel\\Sim7.pdf")


Datafile = "C:\\Users\\jb1203\\Google Drive\\AExeter\\MR\\Clopidogrel\\Simulation2.RData"

set.seed(1234)


Scen        = 6
nSim        = 500
Results     = seResults     = matrix(nrow=nSim,ncol=5)
RES         = seRES         =  matrix(nrow=Scen,ncol=5)
CombRES     = seCombRES      =  matrix(nrow=Scen,ncol=5) 
CombResults = seCombResults = matrix(nrow=nSim,ncol=5)

Beta     = c(-0.5  ,-0.5  ,-0.5 ,-0.5 ,-1   ,-1)
BetaGc   = c(0     ,0     ,0    ,0    ,-0.5 ,-0.5)
ParGT    = c(0     ,0     ,2    ,2    ,2    ,0)
ParGU    = c(0     ,0     ,0    ,0    ,0    ,-0.25)
ParGY    = c(-0.25 ,0     ,0    ,0    ,0    ,-0.1)
ParUT    = c(0     ,0.5   ,0    ,0.5  ,0    ,0.5)

Combine = function(Ests=Ests,SEs=SEs,alpha=0.05){
w           = 1/SEs^2
ws          = w/sum(w)
E           = sum(ws*Ests)
SE          = sqrt(sum((ws*SEs)^2))

E2  = (Ests-E)^2
Q   = sum(w*E2)
df  = length(Ests)-1
Qp  = 1-pchisq(Q,df)
Dec = 0 ; if(Qp>=alpha){Dec=1}

return(list(E=E,SE=SE,Q=Q,p=Qp,Dec=Dec))
}


for(j in 1:Scen){
for(i in 1:nSim){

# Step 1: generate data

n    = 10000
G    = rbinom(n,1,0.3)
U    = rnorm(n) + ParGU[j]*G #+ 3*T
Z    = rnorm(n)
eT   = rnorm(n)
eS   = rnorm(n)

etaT = -3 + ParUT[j]*U + eT + ParGT[j]*G + 0.2*Z
pT   = exp(etaT)/(1+exp(etaT))
T    = rbinom(n,1,pT)


Gc   = 1-G
S    = 0 + Beta[j]*T*G + BetaGc[j]*T*Gc + ParGY[j]*G + 0.5*U + Z + eS


Tstar = T*G

# MR

Tshat = lm(Tstar~G+Z)$fitted
IVfit = lm(S~Tshat+Z)
MR    = summary(IVfit)$coef[2,1]
sMR   = summary(IVfit)$coef[2,2]

# Corrected-As treated (CAT)

ASfit = lm(S~T+Z)
CAT    = summary(ASfit)$coef[2,1]/mean(G[T==1])
sCAT   = summary(ASfit)$coef[2,2]/mean(G[T==1])

# GMTE(1)

FitFD  = lm(S~T+Tstar+Z)
GMTE1     = summary(FitFD)$coef[3,1]
sGMTE1    = summary(FitFD)$coef[3,2]

# GMTE(0)

tt        =  (1-T)
ts        =  tt*G
FitFD     = lm(S~tt+ts+Z)
GMTE0     = summary(FitFD)$coef[3,1]
sGMTE0    = summary(FitFD)$coef[3,2]


# RGMTE

fit    = lm(S~T+Tstar+Tshat+Z) # PERR
RGMTE  = summary(fit)$coef[3,1]
sRGMTE = summary(fit)$coef[3,2]

#######################
# Combined methods    #
#######################

Ests         = c(MR,RGMTE); SEs = c(sMR,sRGMTE)
RGMTE_MR     = Combine(Ests,SEs)
Ests         = c(CAT,RGMTE); SEs = c(sCAT,sRGMTE)
RGMTE_CAT    = Combine(Ests,SEs)
Ests         = c(CAT,MR); SEs = c(sCAT,sMR)
MR_CAT       = Combine(Ests,SEs)
Ests         = c(CAT,GMTE1); SEs = c(sCAT,sGMTE1)
GMTE1_CAT    = Combine(Ests,SEs)
Ests         = c(MR,RGMTE,CAT); SEs = c(sMR,sRGMTE,sCAT)
RGMTE_MR_CAT = Combine(Ests,SEs)


Results[i,]       = c(CAT,GMTE1,GMTE0,RGMTE,MR)
seResults[i,]     = c(sCAT,sGMTE1,sGMTE0,sRGMTE,sMR)
CombResults[i,]   = c(RGMTE_MR$E,RGMTE_CAT$E,MR_CAT$E,GMTE1_CAT$E,RGMTE_MR_CAT$E)
seCombResults[i,] = c(RGMTE_MR$SE,RGMTE_CAT$SE,MR_CAT$SE,GMTE1_CAT$SE,RGMTE_MR_CAT$SE)

print(i)


}


RES[j,]         = round(apply(Results,2,mean),3)
seRES[j,]       = signif(apply(seResults,2,mean),2)
CombRES[j,]     = round(apply(CombResults,2,mean),3)
seCombRES[j,]   = signif(apply(seCombResults,2,mean),2)


D = NULL;for(hh in 1:5){D[hh] = max(density(Results[,hh])$y)+1.5}
D = max(D)


plot(density(Results[,1]),xlim=range(Results[,]),xlab="Distribution of Estimates",
     main="",lwd=4,cex.lab=1.5,col="black",ylim=c(0,D))  # As treated
lines(density(Results[,2]),col="orange",lwd=4)           # GMTE(1)
lines(density(Results[,3]),col="red",lwd=4)              # GMTE0
lines(density(Results[,4]),col="blue",lwd=4)             # RGMTE
lines(density(Results[,5]),col="green",lwd=4,lty=2)      # MR
title(main = paste("Scenario",j),cex.main=3)

if(j==1){legend("topleft",c("CAT",
                    "GMTE(1)",
                    "GMTE(0)",
                    "RGMTE",
                    "MR"),
lwd=4,cex=1.3,col=c("black","orange","red","blue","green"),bty="n")}

lines(rep(-0.5,2),c(0,D),lty=2,lwd=2)

dev.print(device=pdf,file=plotfile[j])
print(j)

}


save(RES,seRES,CombRES,seCombRES,file=Datafile)

Tot = 2*Scen
RESall = matrix(nrow=Tot,ncol=6)
RESall[seq(1,(Tot-1),by=2),2:6] = RES
RESall[seq(2,(Tot),by=2),2:6] = seRES
d          = NULL 
for(i in 1:Scen){d=c(d,rep(i,2))}
RESall[,1] = d


colnames(RESall) = c("Scenario","CAT","GMTE(1)","GMTE0","RGMTE","MR")
rownames(RESall) = rep(c("Est.","S.E"),Scen)


Tot = 2*Scen
CRESall = matrix(nrow=Tot,ncol=6)
CRESall[seq(1,(Tot-1),by=2),2:6] = CombRES
CRESall[seq(2,(Tot),by=2),2:6] = seCombRES
d          = NULL 
for(i in 1:Scen){d=c(d,rep(i,2))}
CRESall[,1] = d

colnames(CRESall) = c("Scenario","RGMTE_MR","RGMTE_CAT","MR_CAT","GMTE1_CAT","RGMTE_MR_CAT")

rownames(CRESall) = rep(c("Est.","S.E"),Scen)

round(RESall,3)
round(CRESall,3)
```
