## Supplementary material for "The Triangulation WIthin A STudy (TWIST) framework for causal inference within Pharmacogenetic research": code: SurvivalDataCode.R

```
Combine = function(Ests=Ests,SEs=SEs,alpha=0.05){
w           = 1/SEs^2
ws          = w/sum(w)
E           = sum(ws*Ests)
SE          = sqrt(sum((ws*SEs)^2))

E2  = (Ests-E)^2
Q   = sum(w*E2)
df  = length(Ests)-1
Qp  = 1-pchisq(Q,df)
Dec = 0 ; if(Qp>=alpha){Dec=1}

P.Est          = 2*(1-pnorm(abs(E/SE)))

R        = c(E,SE,P.Est,Q,Qp,Dec)
ResultsX = matrix(R,nrow=1)
return(ResultsX)
}


	# G = binary genotype
	# T = binary Treatment indicator
	# Survival data.frame (time_0,S_t,S_d) 
      # time_0 = time participants enter
      # S_t is the follow-up time 
      # S_d =  binary variable indicating event or censoring status
      # Z = covariate
	

AalenGMTE = function(G,T,time_0,S_t,S_d,Z){


	require(timereg)

	########################
	# interaction variable #
	########################

	Tstar = T*G

	# IV analysis

	Tstar = T*G 
	Tshat = glm(Tstar~G + Z,family=binomial(link="logit"))$fitted 
	MRfit = fit_aalen = aalen(Surv(time_0, S_t, S_d) ~ const(Tshat) + const(Z), n.sim=100) 
	MR    = coef(MRfit)[1,1] 
	sMR   = coef(MRfit)[1,2] 

      # Corrected-As treated (CAT)

      Tcat   = T*mean(G[T==1]) 
	CATfit = fit_aalen = aalen(Surv(time_0, S_t, S_d) ~ const(Tcat) + const(Z), n.sim=100)
      CAT    = coef(CATfit)[1,1]
	sCAT   = coef(CATfit)[1,2]

	# GMTE(1)

	GMTE1fit  = fit_aalen = aalen(Surv(time_0, S_t, S_d) ~ const(T)+const(Tstar) + const(Z), n.sim=100)
	GMTE1     = coef(GMTE1fit)[2,1]
	sGMTE1    = coef(GMTE1fit)[2,2]

      # GMTE(0)

      tt        =  (1-T)
      ts        =  tt*G
      GMTE0fit  = fit_aalen = aalen(Surv(time_0, S_t, S_d) ~ const(tt)+const(ts) + const(Z), n.sim=100)
	GMTE0     = coef(GMTE0fit)[2,1]
	sGMTE0    = coef(GMTE0fit)[2,2]

	# RGMTE

	RGMTEfit  = fit_aalen = aalen(Surv(time_0, S_t, S_d) ~ const(T)+const(Tstar)+const(Tshat),+ const(Z), n.sim=100) 
	RGMTE     = coef(RGMTEfit)[2,1]
	sRGMTE    = coef(RGMTEfit)[2,2]

#######################
# Combined methods    #
#######################

Ests         = c(MR,RGMTE); SEs = c(sMR,sRGMTE)
RGMTE_MR     = Combine(Ests,SEs)
Ests         = c(CAT,RGMTE); SEs = c(sCAT,sRGMTE)
RGMTE_CAT    = Combine(Ests,SEs)
Ests         = c(CAT,MR); SEs = c(sCAT,sMR)
MR_CAT       = Combine(Ests,SEs)
Ests         = c(CAT,GMTE1); SEs = c(sCAT,sGMTE1)
GMTE1_CAT    = Combine(Ests,SEs)
Ests         = c(MR,RGMTE,CAT); SEs = c(sMR,sRGMTE,sCAT)
RGMTE_MR_CAT = Combine(Ests,SEs)

FullCombined     = matrix(nrow=5,ncol=6)
FullCombined[1,] = RGMTE_MR
FullCombined[2,] = RGMTE_CAT
FullCombined[3,] = MR_CAT
FullCombined[4,] = GMTE1_CAT
FullCombined[5,] = RGMTE_MR_CAT

colnames(FullCombined) =c("Est","SE","P.Est","Qstat","Qp","Combine?")
rownames(FullCombined) =c("RGMTE_MR","RGMTE_CAT","MR_CAT","GMTE1_CAT","RGMTE_MR_CAT")


Results = matrix(nrow=10,ncol=4)

Results[1,] = c(CAT,sCAT,2*(1-pnorm(abs(CAT/sCAT))),1)
Results[2,] = c(GMTE1,sGMTE1,2*(1-pnorm(abs(GMTE1/sGMTE1))),1)
Results[3,] = c(GMTE0,sGMTE0,2*(1-pnorm(abs(GMTE0/sGMTE0))),0)
Results[4,] = c(MR,sMR,2*(1-pnorm(abs(MR/sMR))),1)
Results[5,] = c(RGMTE,sRGMTE, 2*(1-pnorm(abs(RGMTE/sRGMTE))),1)

Results[6:10,] = FullCombined[,c(1,2,3,6)]

Results

colnames(Results) = c("Est","SE","P","Combine")
rownames(Results) = c("CAT","GMTE1","GMTE0","MR","RGMTE",rownames(FullCombined))

ResultsALL = Results


return(list(MRfit=MRfit,CATfit=CATfit,GMTE1fit=GMTE1fit,
       GMTE0fit=GMTE0fit,RGMTEfit=RGMTEfit,FullCombined=FullCombined,
       CAT=CAT,sCAT=sCAT,MR=MR,sMR=sMR,GMTE1=GMTE1,sGMTE1=sGMTE1,
       GMTE0=GMTE0,sGMTE0=sGMTE0,RGMTE=RGMTE,sRGMTE=sRGMTE,ResultsAll=ResultsAll))


}
```
